# Modulation of Gut Microbiota and Gut-Brain Axis as a Therapeutic approach in Traumatic brain injury: Implications for Neurological Outcomes

**DOI:** 10.1101/2025.10.15.25338135

**Authors:** Fatemeh Vosoughian, Zahra Rezanejad, Omid Bahrami, Amir Mohammad kiapasha, Maral Moafi, Aida Mahmoudjanlu, Farbod Tabasi Kakhki, Alireza Zali, Farzan Fahim

## Abstract

**Objective:** This systematic review aimed to evaluate the impact of gut microbiota modulation including probiotics, dietary interventions, and enteral nutrition strategies on neurological recovery, functional outcomes, morbidity, and mortality in patients with traumatic brain injury (TBI).

**Methods:** The review was conducted according to PRISMA 2020 guidelines and registered in PROSPERO. A comprehensive search was performed across PubMed, Embase, Web of Science, Scopus, and Cochrane Library up to September 2025. Inclusion criteria comprised original clinical studies in human TBI patients that investigated microbiota related interventions and reported neurological, gastrointestinal, or microbiome outcomes. Risk of bias was assessed using the Joanna Briggs Institute (JBI) tools.

**Results:** Among five eligible studies (four RCTs and one cohort; n = 725), early or individualized enteral nutrition and probiotic supplementation were generally associated with better metabolic and inflamatory profiles, fewer gastrointestinal complications, and a non significant trend toward improved neurological scores (GCS, GOS). However, effect sizes weres mall and certainty low due to limited sample sizes and methodological heterogeneity.

**Conclusion:** Modulation of the gut brain axis, particularly through early and tailored enteral nutrition, appears to improve clinical outcomes and mitigate complications in TBI patients. Probiotics demonstrate safety and potential benefit, though evidence remains preliminary. Larger, multicenter randomized controlled trials are warranted to confirm efficacy, define optimal timing and formulations, and assess longterm neurological recovery.

## 1. Introduction

Traumatic brain injury (TBI) remains one of the foremost causes of death and long-term disability worldwide, affecting an estimated 64–74 million individuals annually with mortality ranging from 9 to 28 per 100,000 in Europe and exceeding 40 % in low- and middle-income countries (1-3). Beyond the primary mechanical insult, TBI triggers secondary cascades characterized by neuroinflammation, oxidative stress, blood–brain barrier (BBB) disruption, and neuronal apoptosis, which collectively impede neurological recovery (4-6). Despite advances in surgical and pharmacologic management (4, 6), current treatments often fail to attenuate these secondary mechanisms and can introduce further complications such as infection, metabolic derangements, and electrolyte imbalance (7, 8). Therefore, novel adjuvant strategies addressing neuroinflammation and systemic metabolic dysfunction are urgently needed.

Emerging evidence highlights the gut–brain axis (GBA)—a bidirectional network linking intestinal and central nervous systems—as a critical regulator of neuro-immune homeostasis (9-11). The intestinal microbiota modulates immune signaling, produces neuroactive metabolites (e.g., short-chain fatty acids [SCFAs]), and influences BBB integrity (11). Experimental TBI models reveal profound dysbiosis shortly after injury, with loss of beneficial SCFA-producing taxa and increased intestinal permeability that perpetuate systemic and CNS inflammation (12, 13). In a clinical cross-sectional study, Yu et al. (2022) demonstrated that patients with prolonged disorders of consciousness exhibited significantly altered gut compositions and markedly reduced fecal SCFAs—acetic, propionic, and butyric acids—correlating with impaired EEG network connectivity (14). These findings highlight the crucial role of the gut microbiome in post-traumatic neuroplasticity and recovery of consciousness.

On the therapeutic side, nutritional and microbiota-targeted interventions such as early or individualized enteral nutrition (EN), glutamine supplementation, and probiotic therapy are being explored in neurocritical care (15-20). Such interventions may preserve mucosal integrity, attenuate oxidative stress, modulate systemic immunity, and ultimately influence neurological outcomes. In parallel, micronutrient-based strategies, particularly vitamin D and E supplementation, have shown promising neuroprotective potential by mitigating neuroinflammation and oxidative stress. A recent systematic review and meta-analysis demonstrated that vitamin D supplementation significantly improved Glasgow Coma Scale (GCS) scores and functional outcomes following moderate-to-severe TBI (21). Clinical data, albeit limited, support these mechanisms: randomized trials in severe TBI have shown that tailored or early EN protocols reduce infection rates and shorten ICU stay (22, 23), while probiotic supplementation favorably modulates inflammatory markers and adhesion molecules (24, 25). Furthermore, Pan et al. (2025) recently developed a validated nomogram identifying predictors of EN feeding intolerance—mean arterial pressure, antibiotic use, and mechanical ventilation— emphasizing the tight interplay between gastrointestinal function and systemic recovery after TBI (26).

Collectively, these findings highlight the gut–brain–microbiota axis as a promising therapeutic target. However, clinical evidence remains fragmented, with small single-center studies and heterogeneous outcomes. This systematic review, therefore, aims to comprehensively synthesize available clinical data on gut microbiota modulation—including probiotic, prebiotic, and enteral-nutrition-based strategies—in TBI patients, assessing effects on neurological recovery, gastrointestinal tolerance, and overall prognosis. The analysis integrates quantitative meta-evaluation where feasible and narratively summarizes mechanistic insights to elucidate the translational relevance of gut-targeted therapies in TBI management.

## 2. Method

### 2.1. Study Design and Registration

This systematic review was conducted in accordance with the Preferred Reporting Items for Systematic Reviews (PRISMA) 2020 guidelines. The study protocol was developed a priori and registered in the International Prospective Register of Systematic Reviews (PROSPERO; Registration ID: CRD420251159440).

### 2.2. Information Sources and Search Strategy

A comprehensive literature search was conducted across **PubMed (n=219), Embase (n=695), Web of Science (n=377), Scopus (n=1317), and the Cochrane Library (n=19)** untill September 2025. The search strategy combined Medical Subject Headings (MeSH) and free-text terms related to “Traumatic Brain Injury” [MeSH], “Traumatic Brain Injuries”, “Brain Trauma”, “Brain Traumas”, “Traumatic Encephalopathies”, “Traumatic Encephalopathy”, “TBIs”, “TBI”, “Craniocerebral Trauma” [MeSH], “Craniocerebral Traumas”, “Head Injuries”, “Head Injury”, “Craniocerebral Injuries”, “Craniocerebral Injury”, “Head Trauma”, “Head Traumas”, “Frontal Region Trauma”, “Frontal Region Traumas”, “Forehead Trauma”, “Forehead Traumas”, “Occipital Region Trauma”, “Occipital Region Traumas”, “Occipital Trauma”, “Occipital Traumas”, “Parietal Region Trauma”, “Parietal Region Traumas”, “Temporal Region Trauma”, “Temporal Region Traumas”, “Crushing Skull Injury”, “Crushing Skull Injuries”, “Multiple Head Injury”, “Multiple Head Injuries”, “Minor Head Injuries”, “Minor Head Injury”, “Open Head Injuries”, “Open Head Injury”, “Superficial Head Injuries”, “Superficial Head Injury”, “Concussion” [MeSH], “Brain Concussions”, “Cerebral Concussion”, “Cerebral Concussions”, “Commotio Cerebri”, “Intermediate Concussion”, “Intermediate Concussions”, “Mild Concussion”, “Mild Concussions”, “Mild Traumatic Brain Injury”, “Severe Concussion”, “Severe Concussions”, “Gut-Brain Axis” [MeSH], “Brain Gut Axis”, “Brain and Gut Axis”, “Gut and Brain Axis”, “Microbiota-Gut-Brain Axis”, “Microbiota Gut Brain Axis”, “Microbiome-Brain-Gut Axis”, “Microbiome Brain Gut Axis”, “Brain-Gut-Microbiome Axis”, “Brain Gut Microbiome Axis”, “Microbiota-Brain-Gut Axis”, “Microbiota Brain Gut Axis”, “Gut-Brain-Microbiome Axis”, “Gut Brain Microbiome Axis”, “Microbiome-Gut-Brain Axis”, “Microbiome Gut Brain Axis”, “Gastrointestinal Microbiome” [MeSH], “Gastrointestinal Microbiomes”, “Gut Microbiome”, “Gut Microbiomes”, “Gastrointestinal Microbial Community”, “Gastrointestinal Microbial Communities”, “Gut Microflora”, “Gastrointestinal Microflora”, “Gastrointestinal Flora”, “Gut Flora”, “Gastrointestinal Microbiota”, “Gastrointestinal Microbiotas”, “Gut Microbiota”, “Gut Microbiotas”, “Intestinal Microbiome”, “Intestinal Microbiomes”, “Intestinal Microbiota”, “Intestinal Microbiotas”, “Intestinal Flora”, “Intestinal Microflora”, “Enteric Bacteria”, “Gastric Microbiome”, “Gastric Microbiomes”, “Gastrointestinal Complication” [MeSH], “Gastrointestinal Complications”, “Constipation” [MeSH], “Colonic Inertia”, “Dyschezia”, “Diarrhea” [MeSH], “Diarrheas”, “Bacterial Overgrowth” [MeSH], “Small Intestinal Bacterial Overgrowth”, “Dysbiosis” [MeSH], “Dysbioses”, “Dys-symbiosis”, “Dys-symbioses”, “Dys symbiosis”, “Disbiosis”, “Disbioses”, “Dysbacteriosis”, “Disbacterioses”, “Dyspepsia” [MeSH], “Dyspepsias”, “Indigestion”, “Indigestions”, “Weight Loss” [MeSH], “Weight Losses”, “Weight Reduction”, “Weight Reductions”, “Treatment” [MeSH], “Therapeutic”, “Therapy”, “Therapies”, “Treatments”, “Restoration”, “Improvement”, “Management” [MeSH], “Disease-Management”, “Prognosis” [MeSH], “Prognoses”, “Prognostic Factors”, “Prognostic Factor”, “Recovery of Function” [MeSH], “Function Recoveries”, “Function Recovery”.

The search strategy included advanced searches in PubMed, utilizing Boolean operators (AND, OR, NOT), filters for article types, and publication dates to refine and optimize the search results**: ((((((((((((Traumatic Brain injury[MeSH Terms]) OR (Traumatic Brain Injuries[Title/Abstract])) OR (Brain Trauma[Title/Abstract])) OR (Brain Traumas[Title/Abstract])) OR (Traumatic Brain Injury[Title/Abstract])) OR (Traumatic Encephalopathies[Title/Abstract])) OR (Traumatic Encephalopathy[Title/Abstract])) OR (TBIs[Title/Abstract])) OR (TBI[Title/Abstract])) OR ((((((((((((((((((((((((((((((Craniocerebral Trauma[MeSH Terms]) OR (Craniocerebral Traumas[Title/Abstract])) OR (Head Injuries[Title/Abstract])) OR (Head Injury[Title/Abstract])) OR (Craniocerebral Injuries[Title/Abstract])) OR (Craniocerebral Injury[Title/Abstract])) OR (Head Trauma[Title/Abstract])) OR (Head Traumas[Title/Abstract])) OR (Frontal Region Trauma[Title/Abstract])) OR (Frontal Region Traumas[Title/Abstract])) OR (Forehead Trauma[Title/Abstract])) OR (Forehead Traumas[Title/Abstract])) OR (Occipital Region Trauma[Title/Abstract])) OR (Occipital Region Traumas[Title/Abstract])) OR (Occipital Traum[Title/Abstract])) OR (Occipital Traumas[Title/Abstract])) OR (Parietal Region Trauma[Title/Abstract])) OR (Parietal Region Traumas[Title/Abstract])) OR (Temporal Region Trauma[Title/Abstract])) OR (Temporal Region Traumas[Title/Abstract])) OR (Crushing Skull Injury[Title/Abstract])) OR (Crushing Skull Injuries[Title/Abstract])) OR (Multiple Head Injury[Title/Abstract])) OR (Multiple Head Injuries[Title/Abstract])) OR (Minor Head Injuries[Title/Abstract])) OR (Minor Head Injury[Title/Abstract])) OR (Open Head Injuries[Title/Abstract])) OR (Open Head Injury[Title/Abstract])) OR (Superficial Head Injuries[Title/Abstract])) OR (Superficial Head Injury[Title/Abstract]))) OR ((((((((((((CONCUSSION[MeSH Terms]) OR (Brain Concussions[Title/Abstract])) OR (Cerebral Concussion[Title/Abstract])) OR (Cerebral Concussions[Title/Abstract])) OR (Commotio Cerebri[Title/Abstract])) OR (Intermediate Concussion[Title/Abstract])) OR (Intermediate Concussions[Title/Abstract])) OR (Mild Concussion[Title/Abstract])) OR (Mild Concussions[Title/Abstract])) OR (Mild Traumatic Brain Injury[Title/Abstract])) OR (Severe Concussion[Title/Abstract])) OR (Severe Concussions[Title/Abstract]))) AND (((((((((((((((((((((((((Gut brain axis[MeSH Terms]) OR (Brain Gut Axis[Title/Abstract])) OR (Brain[Title/Abstract] AND Gut Axis[Title/Abstract])) OR (Gut[Title/Abstract] AND Brain Axis[Title/Abstract])) OR (Gut-Brain Axis[Title/Abstract])) OR (Microbiota-Gut-Brain Axis[Title/Abstract])) OR (Microbiota Gut Brain Axis[Title/Abstract])) OR (Microbiome-Brain-Gut Axis[Title/Abstract])) OR (Microbiome Brain Gut Axis[Title/Abstract])) OR (Brain-Gut-Microbiome Axis[Title/Abstract])) OR (Brain Gut Microbiome Axis[Title/Abstract])) OR (Microbiota-Brain-Gut Axis[Title/Abstract])) OR (Microbiota Brain Gut Axis[Title/Abstract])) OR (Gut-Brain-Microbiome Axis[Title/Abstract])) OR (Gut Brain Microbiome Axis[Title/Abstract])) OR (Microbiome-Gut-Brain Axis[Title/Abstract])) OR (Microbiome Gut Brain Axis[Title/Abstract])) OR (((((((((((((((((((((((((Gastrointestinal microbiome[MeSH Terms])) OR (Gastrointestinal Microbiomes[Title/Abstract])) OR (GUT microbiome[Title/Abstract])) OR (Gastrointestinal Microbial Community[Title/Abstract])) OR (Gastrointestinal Microbial Communities[Title/Abstract])) OR (Gut Microbiome[Title/Abstract])) OR (Gut Microbiomes[Title/Abstract])) OR (Gut Microflora[Title/Abstract])) OR (Gastrointestinal Microflora[Title/Abstract])) OR (Gastrointestinal Flora[Title/Abstract])) OR (Gut Flora[Title/Abstract])) OR (Gastrointestinal Microbiota[Title/Abstract])) OR (Gastrointestinal Microbiotas[Title/Abstract])) OR (Gut Microbiota[Title/Abstract])) OR (Gut Microbiotas[Title/Abstract])) OR (Intestinal Microbiome[Title/Abstract])) OR (Intestinal Microbiomes[Title/Abstract])) OR (Intestinal Flora[Title/Abstract])) OR (Intestinal Microbiota[Title/Abstract])) OR (Intestinal Microbiotas[Title/Abstract])) OR (Intestinal Microflora[Title/Abstract])) OR (Enteric Bacteria[Title/Abstract])) OR (Gastric Microbiome[Title/Abstract])) OR (Gastric Microbiomes[Title/Abstract]))) OR (Gastrointestinal complication[MeSH Terms]) OR (Gastrointestinal complications [Title/Abstract]))) OR (((Constipation[MeSH Terms]) OR (Colonic Inerti[Title/Abstract])) OR (Dyschezia[Title/Abstract]))) OR ((Diarrhea[MeSH Terms]) OR (Diarrheas[Title/Abstract]))) OR (Bacterial overgrowth[MeSH Terms])OR (small intestinal bacterial overgrowth[Title/Abstract])) OR (((((((((Dysbiosis[MeSH Terms]) OR (Dysbioses[Title/Abstract])) OR (Dys-symbiosis[Title/Abstract])) OR (Dys-symbioses[Title/Abstract])) OR (Dys symbiosis[Title/Abstract])) OR (Disbiosis[Title/Abstract])) OR (Disbioses[Title/Abstract])) OR (Dysbacteriosi[Title/Abstract])) OR (Disbacterioses[Title/Abstract]))) OR ((((Dyspepsia[MeSH Terms]) OR (Dyspepsias[Title/Abstract])) OR (Indigestion[Title/Abstract])) OR (Indigestions[Title/Abstract]))) OR ((((Weight loss[MeSH Terms]) OR (Weight Losses[Title/Abstract])) OR (Weight Reduction[Title/Abstract])) OR (Weight Reductions[Title/Abstract])))) AND ((((((((((Treatment[MeSH Terms]) OR (Therapeutic[Title/Abstract])) OR (Therapy[Title/Abstract])) OR (Therapies[Title/Abstract])) OR (Treatments[Title/Abstract])) OR (Restoration[Title/Abstract])) OR (Improvement[Title/Abstract])) OR ((Management[MeSH Terms]) OR (Disease-Management[Title/Abstract]))) OR ((((Prognosis[MeSH Terms]) OR (Prognoses[Title/Abstract])) OR (Prognostic Factors[Title/Abstract])) OR (Prognostic Factor[Title/Abstract]))) OR (((Recovery of function[MeSH Terms]) OR (Function Recoveries[Title/Abstract])) OR (Function Recovery[Title/Abstract])))**.

### 2.3. Eligibility Criteria

#### Studies were included if they

- Enrolled human participants diagnosed with traumatic brain injury;
- Investigated gut microbiome composition, alterations, or interventions targeting microbiota (e.g., probiotics, prebiotics, fecal microbiota transplantation, dietary modulation);
- Studies that implemented or assessed interventions targeting the Gut–Brain Axis (GBA) and/or gut microbiome.
- Reported clinical or functional outcomes in TBI patients (e.g., Glasgow Coma Scale [GCS], Glasgow Outcome Scale [GOS/GOS-E], mortality, hospital/ICU length of stay, infection rates, inflammatory or metabolic biomarkers);
- Any original human studies (randomized/non-randomized trials, cohort, case–control, cross-sectional, before–after, registry studies, case series ≥5 patients)
- Studies enrolling patients with Traumatic Brain Injury (TBI) (mild/moderate/severe; acute, subacute, or chronic phases).
- Articles that reported at least one neurological or gastrointestinal outcome.
- No language limitation (studies in any language).

#### Studies were excluded if they

- Animal studies, in vitro investigations, reviews, meta-analyses, case reports, conference abstracts, protocols, preprints, letters, and case series with <5 patients;
- Lacked relevant or extractable clinical outcome measures.
- Studies where the intervention does not target the GBA/microbiome (e.g., vitamins/minerals or neuroprotective drugs without a gut/microbiome component).
- Articles without relevant outcomes (neither neurological nor GI nor microbiome/GBA outcomes), or insufficient data for extraction.
- Pediatric or neonatal populations
- Studies not involving TBI patients or Studies that combine TBI data with other brain injuries (e.g., stroke, subarachnoid hemorrhage) without reporting separate subgroup analyses for TBI.
- Studies reporting only microbiome alterations without linking them to neurological outcomes or TBI-specific effects.

### 2.4. Study Selection

The full search strategy for other databases is provided in Supplementary Table S1. All retrieved records were exported into **EndNote** for initial deduplication (999 records removed). The remaining records were imported into **Rayyan**, where an additional 176 duplicates were removed. After de-duplication, **1628 records** were screened. Each article was independently screened by two reviewers. Title/abstract screening was independently performed by O.B., M.M., A.M., and Conflicts were resolved by F.V. Following title and abstract screening, Full texts of 91 studies were retrieved and assessed independently by O.B. and Z.R. according to the inclusion and exclusion criteria. Disagreements were resolved by discussion or, if necessary, adjudication by a third reviewer (F.V.). The remaining 1537 records were excluded due to the following reasons: animal studies (n = 345), irrelevant or unsuitable outcomes (n = 567), case reports (n = 181), case series (n = 26), reviews and meta-analyses (n = 356), in vitro studies (n = 49), and inappropriate study types such as protocol studies (n = 8), cross-sectional studies (n = 4), and conference abstracts (n = 1). Eventually, **five** studies met the predefined inclusion criteria and were included in the final analysis. Among these, 1 was a cohort study and 4 were randomized controlled trials (RCTs). The study selection process is summarized in a PRISMA flow diagram (Figure 1).

**Figure 1.**
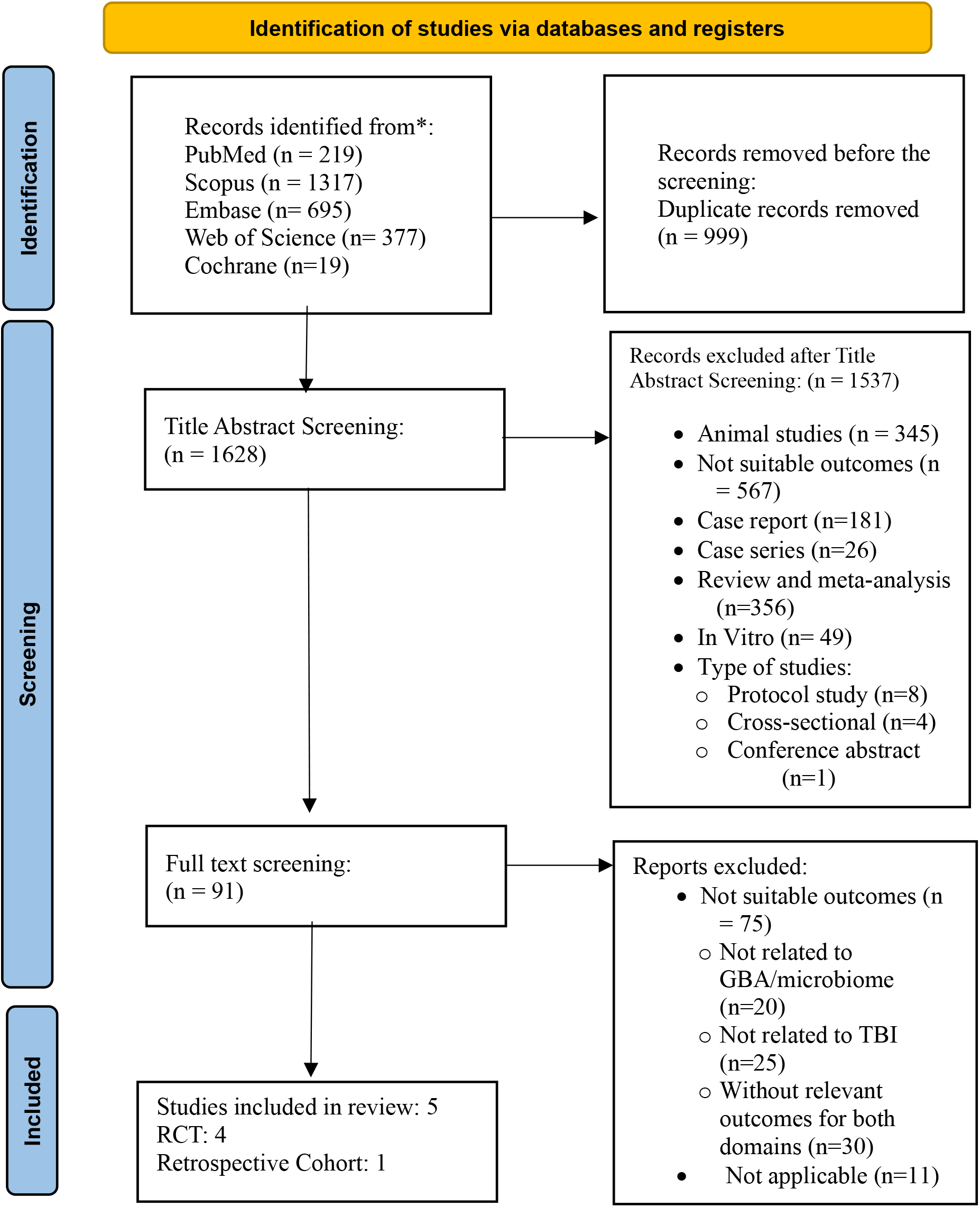
PRISMA Flow chart

### 2.5. Data Extraction

5 studies met the inclusion criteria and were retained for final analysis. Among these, 1 was a cohort study and 4 were randomized controlled trials (RCTs). Data were independently extracted by O.B. and Z.R. using a standardized pilot-tested form. Extracted variables included: first author, country, funding source/conflict of interest, study design, effect size, bias selection/ confounding : matching or regression, Setting, Population (age, sex, undelying disease), TBI severity, Time from injury to intervention, intervention, comparator, Concomitant Treatments, Sample Size intervention, Sample Size control, Loss to Follow-up / Attrition (number of withdrawals/dropouts, Sex distribution of withdrawals, Reason for withdrawal), Baseline characteristics (p value)(Age, male (n), GCS score on admission mean, Time from injury to ICU mean), Follow-up Duration, Primary Outcomes, Secondary Outcomes, Microbiome Outcome Details (diversity (Alpha diversity, Beta diversity), composition, F/B ratio), Outcomes Measurement Tools, Effect size (CI) (Binary, Continuous, Time to event), Statistical Adjustments / Covariates (Adjusted, unadjusted), Adverse Events (Occurrence / Reported (Yes/No / Not reported), Type of adverse event, Frequency in intervention group), Microbiome Assessment Method (sample, Processing, Sequencing method), Main Findings, Risk of Bias, GRADE Assessment, Notes. Any discrepancies were resolved by consensus or through a third reviewer (F.V.). The extracted data from the included studies are presented in Supplementary Table S2, which provides a comprehensive summary of their key characteristics.

### 2.6. Risk of Bias Assessment

Each included study was independently assessed for risk of bias by F.V. and F.T. using the appropriate Joanna Briggs Institute (JBI) Critical Appraisal Checklist: the JBI checklist for randomized controlled trials for RCTs and the JBI checklist for cohort studies for cohort designs. Disagreements were resolved through consensus or a third reviewer. No study was excluded based on high risk of bias. The risk of bias assessments are summarized in Table 1.

**Table 1.** The risk of bias assessment JBI Critical Appraisal checklists.

| *Study* | *Num of positive answers* | *Num of negative answers* | *Num of partial/unclear answers* | *Final status* |
| --- | --- | --- | --- | --- |
| ***Randomized control trial studies*** |  |  |  |  |
| *Zhu et al. 2021* | 9 | - | 4 | Moderate |
| *Brenner et al. 2020* | 9 | - | 4 | Moderate |
| *Wang et al. 2023* | 9 | - | 4 | Moderate |
| *Xiong et al. 2021* | 10 | 2 | 1 | High |
| ***Cohort studies*** |  |  |  |  |
| *Pan et al. 2025* | 9 | 2 | - | Moderate |

### 2.7. Certainty of evidence (GRADE)

The overall certainty of evidence for each outcome was assessed using the GRADE (G rading of R ecommendations, A ssessment, D evelopment, and E valuation) approach, considering risk of bias, inconsistency, indirectness, imprecision, and publication bias.

### 2.8. Eligibility for quantitative synthesis

Following completion of the systematic review data extraction from all eligible studies, individual outcome domains were assessed for suitability for quantitative synthesis. Continuous neurological outcomes, including Glasgow Coma Scale (GCS) scores, and binary gastrointestinal tolerance outcomes (feeding intolerance incidence) were pre specified in the protocol as meta analysable endpoints, conditional on ≥2 randomized controlled trials (RCTs) reporting sufficient comparable data. For GCS, two RCTs (Wang 2023 (n = 152) and Zhu 2021 (n = 80)) provided group means, standard deviations (SD), and sample sizes. For feeding intolerance, two RCTs)Zhu 2021 and Xiong 2021 (n = 133 combined)) reported incidence data as number of events over total participants per arm.

### 2.9. Data extraction and preparation

GCS scores were abstracted as post intervention mean ± SD for intervention and control arms at the shortest reported follow up (day 14 for Zhu; day 14 for Wang), consistent with the acute/subacute recovery period following TBI. Feeding intolerance was defined in both trials as the occurrence of one or more gastrointestinal adverse events during early ICU feeding (abdominal distension, diarrhea, constipation, gastric retention, or refeeding syndrome); event counts (n) and totals (N) per arm were extracted exactly as reported in trial tables. All numerical data were verified against published full texts and cross checked with the original extraction spreadsheet (Data extraction FOR TBI AND NUTRITION.xlsx, sheets corresponding to study IDs “33” and “40” for Zhu 2021 and Xiong 2021; “18” for Wang 2023).

### 2.10. Statistical analysis

Meta-analyses were performed using R (version 4.5.1; meta and metafor packages). For continuous outcomes (e.g., Glasgow Coma Scale), standardized mean differences (SMDs) with 95 % confidence intervals were computed using the HartungKnapp adjustment for random effects models. For binary outcomes (e.g., feeding intolerance, infection), risk ratios (RRs) were pooled using the same model. Between-study heterogeneity was quantified with I^2^ and τ^2^. GRADE methods were applied to rate the certainty of evidence for each outcome domain.

A random effects model was applied for all pooled effect estimates, using the inverse variance method for weighting and the Hartung–Knapp–Sidik–Jonkman adjustment to improve small sample inference. Statistical heterogeneity was quantified with the χ^2^ test (Cochran’s Q), the I^2^ statistic (percentage of total variability attributable to between study differences), and τ^2^ (between study variance). Where appropriate, a 95% prediction interval was calculated to convey the range of likely true effects in future comparable studies. Forest plots were generated with modern syntax avoiding deprecated arguments, specifying left hand study and event columns, diamond markers for pooled effects, and green squares for study level estimates. Funnel plots were visually inspected to assess potential small study publication bias; however, with only two studies per outcome, Egger’s regression test was not performed due to low power.

### 2.11. Handling of potential biases and sensitivity

Given the small number of included trials per outcome, sensitivity analyses were limited to fixed effect models for comparison and exclusion of high-risk-of-bias studies. Risk of bias was assessed via the JBI. Notably, both feeding intolerance trials were single-center and only one was single-blinded (Xiong 2021); for GCS, neither trial applied blinding. No imputation for missing data was performed, as all required numerical inputs were available.

### 2.12. Outcome metrics reported

For GCS, the pooled MD with 95% CI, p value, I^2^%, τ^2^, and prediction interval were reported. For feeding intolerance, pooled RR with 95% CI, p value, I^2^%, τ^2^, and prediction interval were reported alongside absolute event rates by arm for each study. Directionality was maintained such that values <1.0 for RR indicate reduced risk of intolerance with intervention feeding, and positive MD values for GCS indicate improved neurological function in the intervention arm.

### 2.13. Outcomes

#### 2.13.1. Primary outcome

- The effect of gut microbiome alterations on neurological recovery in TBI patients, assessed by the Glasgow Coma Scale (GCS) and the Glasgow Outcome Scale (GOS/GOS-E)

#### 2.13.2. Secondary outcomes

- Glasgow Outcome Scale (GOS/GOS-E);
- Mortality;
- Duration of ICU stay, hospital stay, and mechanical ventilation;
- Incidence of infections and sepsis;
- Inflammatory, metabolic, and immune biomarkers;
- Safety and adverse events related to microbiome interventions.

### 2.14. Ethics

No ethics approval was required for this systematic review, as only previously published, deidentified data were used.

## 3. Result

### 3.1. Study selection

A total of 2627 records were identified through database searching. After removal of 999 duplicates, 1628 unique articles were screened by title and abstract using Rayyan. Of these, 91 articles were assessed in full text, and 5 studies were included in the final analysis (Figure 1).

### 3.2. Risk of Bias assessment

Each included study was independently assessed for risk of bias by two reviewers using the appropriate Joanna Briggs Institute (JBI) Critical Appraisal Checklist: the JBI checklist for randomized controlled trials for RCTs and the JBI checklist for cohort studies for cohort designs. Disagreements were resolved through consensus or a third reviewer. No study was excluded based on a high risk of bias. The risk of bias assessments is summarized in Table 1.

### 3.3. Meta-analysis

#### 3.3.1. GCS outcome

Two RCTs comprising 232 participants (117 in the intervention group, 115 in the control group) were included in the GCS meta-analysis (22, 23).

- Wang 2023: Early EN group had a mean GCS of 8.60 ± 1.30 vs 8.80 ± 1.40 in delayed EN (Mean Difference [MD] = –0.20, p = 0.366).
- Zhu 2021: Individualized EN group had a mean GCS of 6.46 ± 1.21 vs 6.75 ± 2.07 in conventional EN (MD = –0.29, p = 0.45).

The pooled Standardized Mean Difference under a random-effects model was:

- SMD = –0.15 (95% CI: –0.29 to –0.02)
- p-value (overall effect) ≈ 0.025
- I^2^ = 0% (τ^2^ = 0.00, p for heterogeneity = 0.9366)

Forest plot interpretation:

When results from both studies were combined, only a minimal difference in GCS was observed between early or individualized feeding and standard care (pooled SMD = –0.15 [–0.29 to – 0.02], I^2^ = 0 %), equivalent to roughly 0.2 to 0.3 points on the GCS scale. Although this reached statistical significance, it does not represent a clinically meaningful improvement in consciousness. Rather than indicating neurological decline, the small negative mean reflects variation across studies with similar baseline severity and should be interpreted as no meaningful short-term change in GCS (Figure 2).

**Figure 2.**
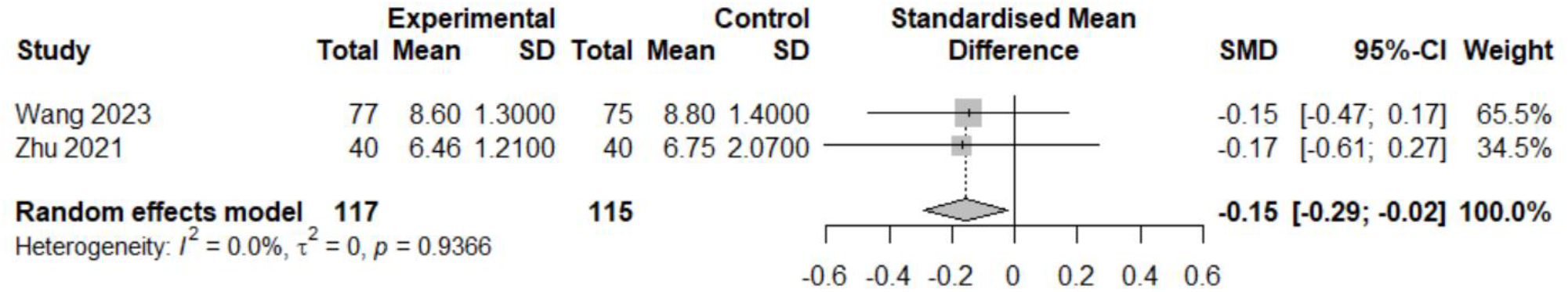
Forest plot for GCS outcome

#### 3.3.2. Feeding Intolerance

Two randomized controlled trials from China (Zhu 2021 and Xiong 2021) enrolling a total of 133 patients with severe traumatic brain injury contributed data on the occurrence of gastrointestinal intolerance to enteral feeding (23, 27). Both trials defined *feeding intolerance* as the occurrence of at least one adverse gastrointestinal event such as abdominal distension, diarrhea, constipation, gastric retention, or refeeding syndrome, recorded during the early phase of ICU admission.

In Zhu 2021, individualized enteral nutrition formulas tailored to digestive function, comorbidities, and metabolic requirements resulted in a 10% incidence of intolerance (4/40) versus 27.5% (11/40) in the standard protocol group (RR = 0.36, 95% CI 0.13–1.05) (23).

In Xiong 2021, a regimen of low-protein, hypocaloric nutrition supplemented with glutamine for seven days was associated with a 33% intolerance rate (9/27) compared to 69.2% (18/26) in the full-feeding control group (RR = 0.48, 95% CI 0.27–0.87) (27).

Pooled analysis using a Hartung-Knapp–adjusted random-effects model indicated a relative risk of 0.45 (95% CI 0.10–2.06), with no statistical heterogeneity (I^2^ = 0%, p = 0.65). The prediction interval was wide (0.02–12.80), reflecting substantial uncertainty from the small number of studies and participants. The combined effect estimate suggests a potential 55% relative reduction in risk; however, the effect did not achieve conventional statistical significance at the 0.05 level (p ≈ 0.095) (Figure 3).

**Figure 3.**
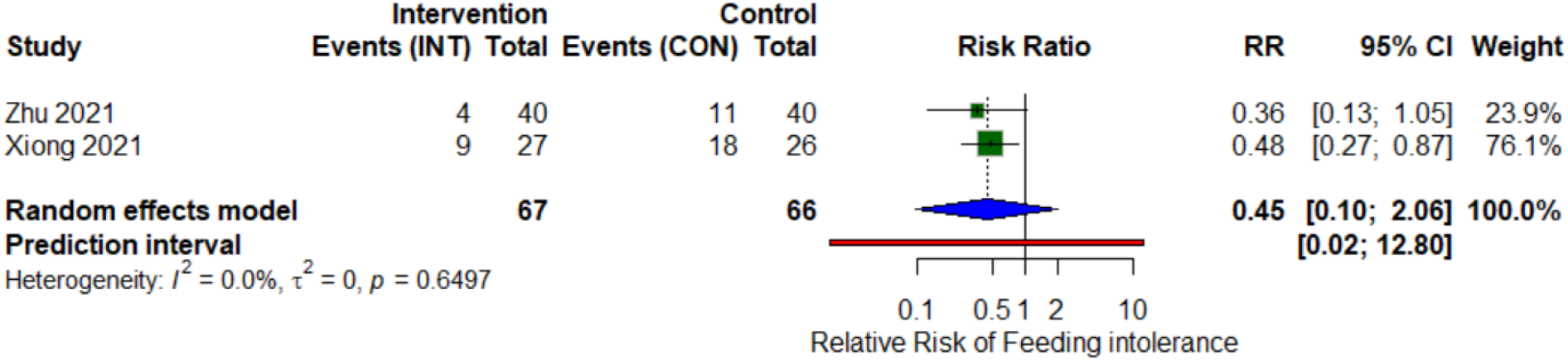
Forest plot for GCS outcome

### 3.3. Study characteristics

This systematic review included five studies: four randomized controlled trials and one retrospective cohort study, published between 2020 and 2025 (22, 23, 26-28). Four studies were conducted in China (22, 23, 26, 27), while one pilot trial originated from the United States (28). Two studies (26, 28), reported external funding; the remaining studies did not disclose financial support.

Sample Sizes ranged from 31 to 409, totaling 725 participants. The majority of participants were male, with proportions ranging from 56.6% (22) to 100% (28). Follow-up durations ranged from 5 days to 3 months. Participant ages ranged from approximately 37 years, young veterans in (28) to 60 years, older ICU patients in (26).

Four studies reported Glasgow Coma Scale (GCS) scores ranging from 3 to 13, encompassing mild to severe TBI (22, 23, 26, 27). Most participants were classified within the moderate to severe range (GCS 3–8). One study focused exclusively on veterans with mild TBI; GCS scores were not explicitly stated (summarized in table 2,4).

**Table 2.** Basic Characteristics and sample populations of eligible studies. Abbreviations: RCT, randomized controlled trial; TBI, traumatic brain injury; GCS, Glasgow Coma Scale; NR, not reported; ICU, intensive care unit; SD, standard deviation

| Study | Study design | Sample size (Intervention/Control) | Participants Age (mean±SD) | Gender (Male/Female %) | TBI severity | Time from injury to intervention | Time from injury to ICU | Follow-up duration | Loss to follow up | fund |
| --- | --- | --- | --- | --- | --- | --- | --- | --- | --- | --- |
| Pan Y et al. (26) | Retrospective cohort | (302/107) | 59.63 ± 13.50 | 66.6/33.4 | Severe, GCS: 3-8 | 24 h | NR | 5 days | NR | yes |
| Wang et al. (22) | RCT | (77/75) | 45.93 ± 9.64 | 56.6/43.4 | Moderate, severe, GCS <13 | ≤48h | NR | 2weeks to 3 months | Yes (4 participants) | no |
| Zhu C et al. (23) | RCT | (40/40) | 46.03 ± 14.65 | 60/40 | severe, moderate, GCS ≤ 8 | NR | NR | 1,7,14 days after intervention | NR | no |
| Xiong W et al. (27) | RCT | (27/26) | 48.88 ± 12.90 | 69.8/30.2 | severe, moderate, GCS: 4 - 8 | within 24 hrs of ICU admission | <8 hrs | 28 days | NR | no |
| Brenner et al. (28) | RCT | (16/15) | 37.40 ± 6.70 | 100/0 | Mild TBI, GCS =NR | Median 10.5 years | NR | 8 ± 2 weeks | Yes (4 participants) | yes |

All studies evaluated nutritional interventions in TBI management, employing distinct strategies. Wang et al. (22) implemented early enteral nutrition (EN) within 48 hours post-injury. Zu et al. (23) utilized individualized EN formulations. Xiong et al. (27) administered a low-protein, hypocaloric EN enriched with glutamine. Pan et al. examined tolerance to standard EN protocols. Four studies delivered EN via nasogastric or nasoenteric tubes. In contrast, Brenner et al. (28) investigated oral probiotic supplementation using Lactobacillus reuteri DSM 17938 in patients with mild TBI. (See Table 3).

**Table 3.**
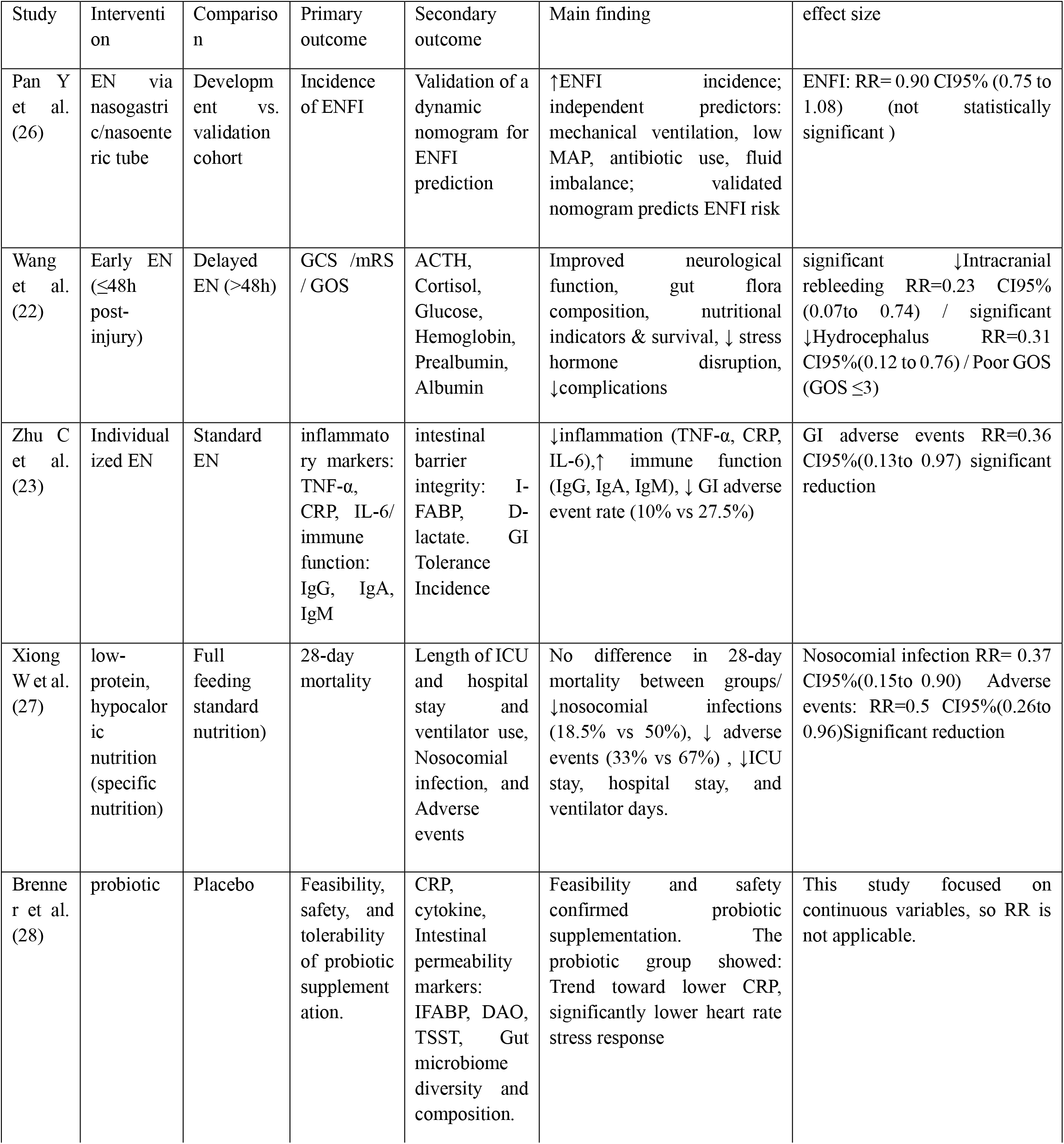
Designs, interventions, outcomes and effect sizes of studies. Abbreviations: EN = Enteral Nutrition; ENFI = Enteral Nutrition Feeding Intolerance; GCS = Glasgow Coma Scale; GOS = Glasgow Outcome Scale; mRS = Modified Rankin Scale; CRP = C-reactive protein; I-FABP = Intestinal Fatty Acid Binding Protein; DAO = Diamine Oxidase; TSST = Trier Social Stress Test; RR = Relative Risk; CI = Confidence Interval.

| Study | Intervention | Comparison | Primary outcome | Secondary outcome | Main finding | effect size |
| --- | --- | --- | --- | --- | --- | --- |
| Pan Y et al. (26) | EN via nasogastric/nasointestinal tube | Development vs. validation cohort | Incidence of ENFI | Validation of a dynamic nomogram for ENFI prediction | ↑ENFI incidence; independent predictors: mechanical ventilation, low MAP, antibiotic use, fluid imbalance; validated nomogram predicts ENFI risk | ENFI: RR= 0.90 CI95% (0.75 to 1.08) (not statistically significant) |
| Wang et al. (22) | Early EN (≤48h post-injury) | Delayed EN (>48h) | GCS /mRS / GOS | ACTH, Cortisol, Glucose, Hemoglobin, Prealbumin, Albumin | Improved neurological function, gut flora composition, nutritional indicators & survival, ↓ stress hormone disruption, ↓ complications | significant ↓Intracranial rebleeding RR=0.23 CI95% (0.07to 0.74) / significant ↓Hydrocephalus RR=0.31 CI95%(0.12 to 0.76) / Poor GOS (GOS ≤3) |
| Zhu C et al. (23) | Individualized EN | Standard EN | inflammatory markers: TNF-α, CRP, IL-6/ immune function: IgG, IgA, IgM | intestinal barrier integrity: I-FABP, D-lactate. GI Tolerance Incidence | ↓inflammation (TNF-α, CRP, IL-6), ↑ immune function (IgG, IgA, IgM), ↓ GI adverse event rate (10% vs 27.5%) | GI adverse events RR=0.36 CI95%(0.13to 0.97) significant reduction |
| Xiong W et al. (27) | low-protein, hypocaloric nutrition (specific nutrition) | Full feeding standard nutrition) | 28-day mortality | Length of ICU and hospital stay and ventilator use, Nosocomial infection, and Adverse events | No difference in 28-day mortality between groups/ ↓nosocomial infections (18.5% vs 50%), ↓ adverse events (33% vs 67%) , ↓ICU stay, hospital stay, and ventilator days. | Nosocomial infection RR= 0.37 CI95%(0.15to 0.90) Adverse events: RR=0.5 CI95%(0.26to 0.96)Significant reduction |
| Brenner et al. (28) | probiotic | Placebo | Feasibility, safety, and tolerability of probiotic supplementation. | CRP, cytokine, Intestinal permeability markers: IFABP, DAO, TSST, Gut microbiome diversity and composition. | Feasibility and safety confirmed probiotic supplementation. The probiotic group showed: Trend toward lower CRP, significantly lower heart rate stress response | This study focused on continuous variables, so RR is not applicable. |

**Table 4.** GCS on Admission: Intervention vs. Control Groups. NA=not applicable.

| Study | intervention Group GCS | Control Group GCS | p-value |
| --- | --- | --- | --- |
| Pan Y et al. (26) | GCS 3–5:(57.62%)/ GCS 6–8: (42.38%) | GCS 3–5: (67.29%)/ GCS 6–8: (32.71%) | 0.101 |
| Wang et al. (22) | Mean GCS: $6.6 \pm 2.5$ | Mean GCS: $6.5 \pm 2.4$ | 0.82 |
| Zhu C et al. (23) | Mean GCS: $6.46 \pm 1.21$ | Mean GCS: $6.42 \pm 1.18$ | 0.78 |
| Xiong W et al. (27) | GCS 4: 4 (14.8%)/<br>GCS 5: 5 (18.6%)/<br>GCS 6: 12 (44.4%)/<br>GCS 7: 4 (14.8%)/<br>GCS 8: 2 (7.4%) | GCS 4: 3 (11.5%)/<br>GCS 5: 8 (30.8%)/<br>GCS 6: 4 (15.4%)/<br>GCS 7: 6 (23.1%)/<br>GCS 8: 5 (19.2%) | 0.163 |
| Brenner et al. (28) | Not reported (mild TBI inferred GCS 13–15) | Not reported | NA |

Primary outcomes assessed across studies included neurological status (GCS, Glasgow Outcome Scale [GOS]), inflammatory markers, morbidity, and mortality.

### 3.4. Neurological outcome

#### 3.4.1. GCS

Baseline GCS scores were reported in all four acute-phase studies and served as indicators of initial neurological status before intervention. Across these studies, intervention and control groups exhibited comparable GCS distributions, suggesting balanced injury severity and minimizing baseline bias.

Pan et al. (26) (total n = 409) investigated the incidence of enteral nutrition feeding intolerance (ENFI) using tube-based enteral nutrition administered via nasogastric or nasoenteric routes. The study compared a development cohort (intervention group, n = 302) with a validation cohort (control group, n = 107). Patients were stratified into two severity categories: GCS 3–5 and GCS 6–8. In the intervention group, 57.62% of patients presented with GCS scores between 3 and 5, while 42.38% fell within the 6–8 range. The control group showed a similar distribution, with 67.29% in the 3–5 range and 32.71% in the 6–8 range. The difference was not statistically significant (p = 0.101), indicating comparable baseline severity.

Wang et al. (22) (total n =152) evaluated the effects of early enteral nutrition initiated within 48 hours post-injury, compared to delayed enteral nutrition initiated after 48 hours. The intervention group included 77patients, and the control group included 75 patients. Mean GCS scores were 6.6 ± 2.5 in the intervention group and 6.5 ± 2.4 in the control group (p = 0.82), demonstrating no significant difference at baseline.

Zhu et al. (23) (total n = 80) implemented individualized enteral nutrition formulations tailored to patient-specific metabolic and clinical needs, compared to standard enteral nutrition. The intervention group comprised 40 patients, and the control group also comprised 40 patients. Mean admission GCS scores were 6.46 ± 1.21 in the intervention group and 6.42 ± 1.18 in the control group (p = 0.78), indicating balanced neurological status at baseline.

Xiong et al. (27) (total n = 31) administered a low-protein, hypocaloric enteral nutrition enriched with glutamine, compared to standard full-feeding enteral nutrition. The intervention group included 16 patients, and the control group included15 patients. In the intervention group, GCS 6 was most prevalent (44.4%), followed by GCS 5 (18.6%), GCS 4 and 7 (14.8% each), and GCS 8 (7.4%). In contrast, the control group exhibited a different distribution: GCS 5 (30.8%), GCS 7 (23.1%), GCS 8 (19.2%), GCS 6 (15.4%), and GCS 4 (11.5%). Statistical analysis revealed no significant difference in GCS distribution between groups (p = 0.163).

Brenner et al. (28) (total n = 31) investigated the feasibility and safety of oral probiotic supplementation using Lactobacillus reuteri DSM 17938 in patients with chronic-phase mild TBI, compared to placebo. The intervention group included 16 participants, and the control group included 15 participants. GCS scores were not reported, as the assessment was not applicable to the study population.

Overall, the absence of statistically significant differences in baseline GCS scores across studies supports the validity of post-intervention comparisons and enhances the internal consistency of the review (see Table 2,3,4).

#### 3.4.2. GOS

Among the included studies, only Wang et al. (22) explicitly evaluated neurological outcomes using the Glasgow Outcome Scale (GOS). In this randomized controlled trial of 152 patients (77 in the early EN group, 75 in the delayed EN group) with TBI (mean GCS score at admission was 6.6 ± 2.5), early EN initiated within 48 hours post-injury was associated with improved neurological recovery. Patients with GCS ≤8 had a significantly higher risk of poor outcomes, defined as GOS ≤3, with a relative risk (RR) of 2.45 (95% CI: 1.42–4.23), and elevated mortality (RR = 3.12, 95% CI: 1.58–6.17). Conversely, patients with GCS >8 had a lower risk of poor outcomes (RR = 0.41, 95% CI: 0.24–0.70) and reduced mortality (RR = 0.32, 95% CI: 0.16–0.64). Although other studies described neurological improvement or stability, none explicitly reported GOS scores, therefore were excluded from GOS-specific analysis (table 5).

**Table 5.**
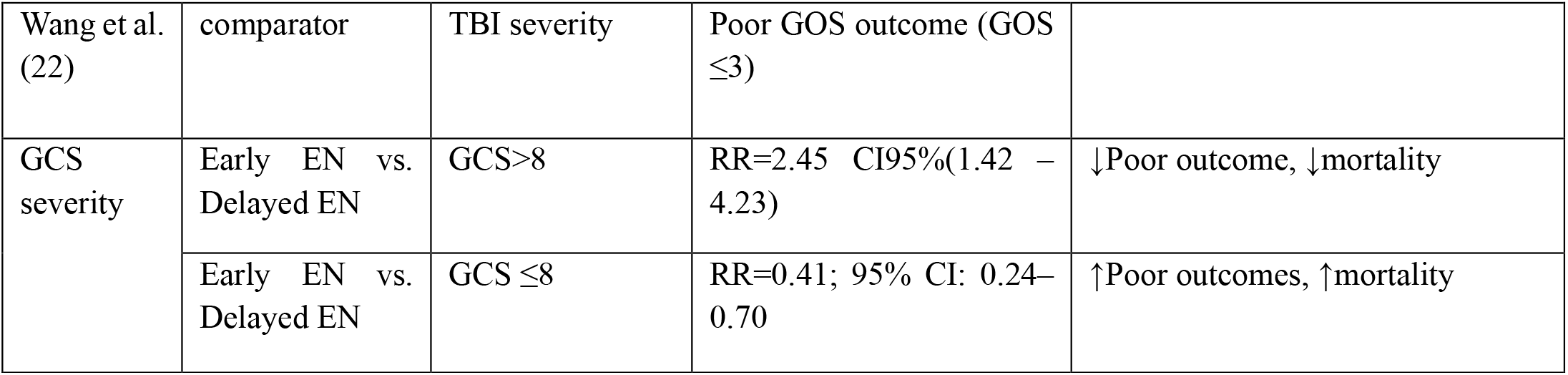
GCS and GOS outcome of wang et al.2023.

|  |  |  |  |  |
| --- | --- | --- | --- | --- |
| Wang et al. (22) | comparator | TBI severity | Poor GOS outcome (GOS $\leq 3$) | |
| GCS severity | Early EN vs. Delayed EN | $GCS > 8$ | RR=2.45 CI95%(1.42 – 4.23) | ↓Poor outcome, ↓mortality |
| | Early EN vs. Delayed EN | $GCS \leq 8$ | RR=0.41; 95% CI: 0.24–0.70 | ↑Poor outcomes, ↑mortality |

### 3.5. Clinical outcomes

#### 3.5.1 Morbidity

Morbidity outcomes varied across studies based on the EN strategy used. Four studies (22, 23, 27, 28) found statistically significant reductions in morbidity outcomes with tailored EN approaches. Gastrointestinal intolerance—including diarrhea, vomiting, and gastric retention—was significantly reduced in intervention groups. Nosocomial infections were reduced in studies with early EN and individualized formula interventions.

Zhu et al. (23) reported improved gastrointestinal tolerance with individualized EN, reflected in a reduced rate of GI adverse events (RR = 0.36, 95% CI: 0.13–0.97). Similarly, Xiong et al. (27) found that glutamine-supplemented EN significantly lowered nosocomial infection rates (RR = 0.37, 95% CI: 0.15–0.90) and general adverse events such as vomiting (RR = 0.50, 95% CI: 0.26– 0.96). Wang et al. (22) demonstrated that early EN was associated with a marked decrease in neurological complications, including intracranial rebleeding (RR = 0.23, 95% CI: 0.07–0.74) and hydrocephalus (RR = 0.31, 95% CI: 0.12–0.76). Pan Y et al. (26) developed a nomogram to predict EN feeding intolerance (ENFI), but found no statistically significant difference between cohorts (RR = 0.90, 95% CI: 0.75–1.08). Although Brenner et al. (28) did not report RR values, their pilot study indicated that probiotic supplementation was safe and feasible, with reduced stress-related physiological responses. These findings support the hypothesis that early and individualized EN improves clinical outcomes in TBI patients by reducing complications like infections, GI intolerance, and neurological risks (Table 6).

**Table 6.**
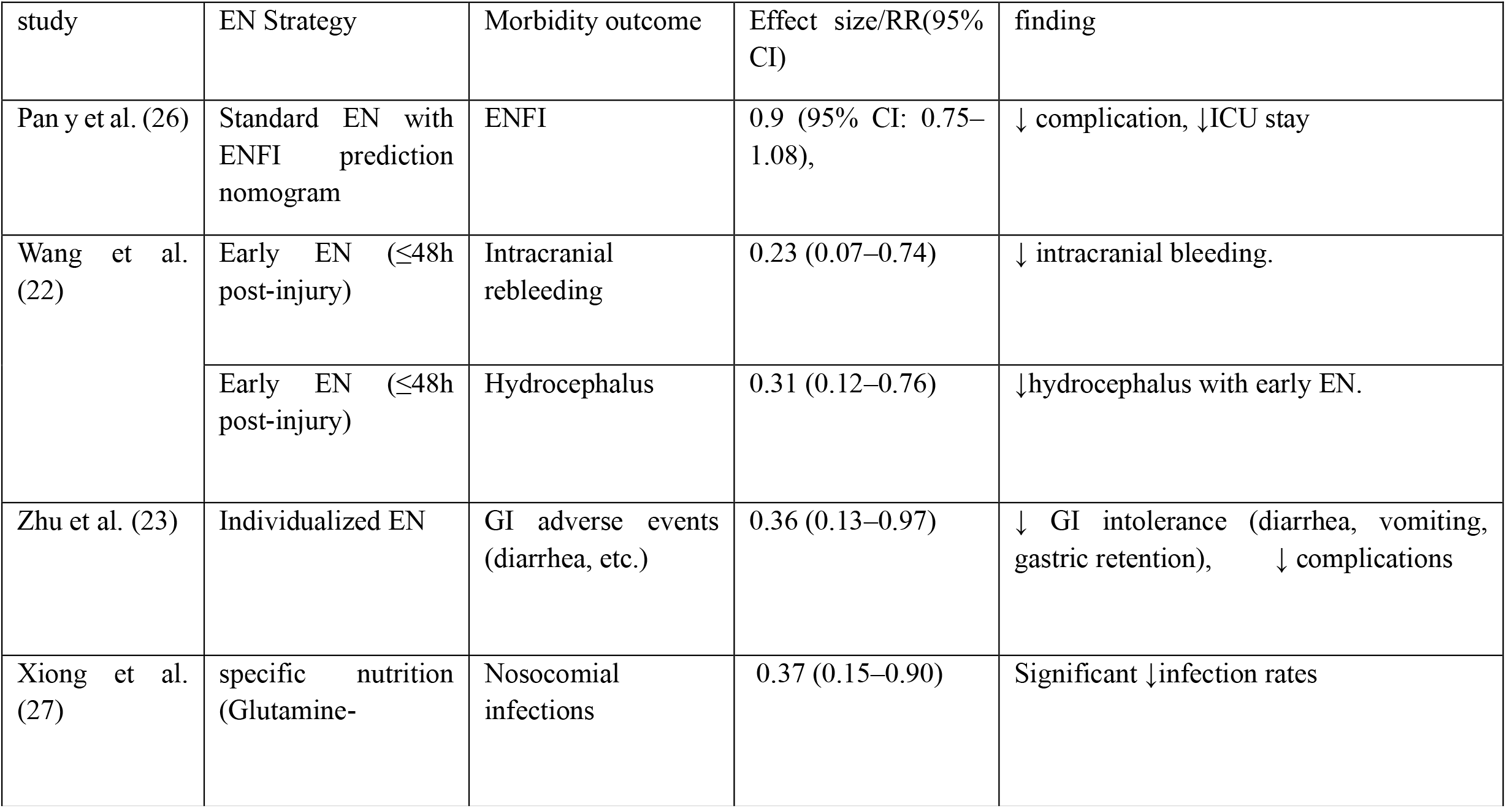
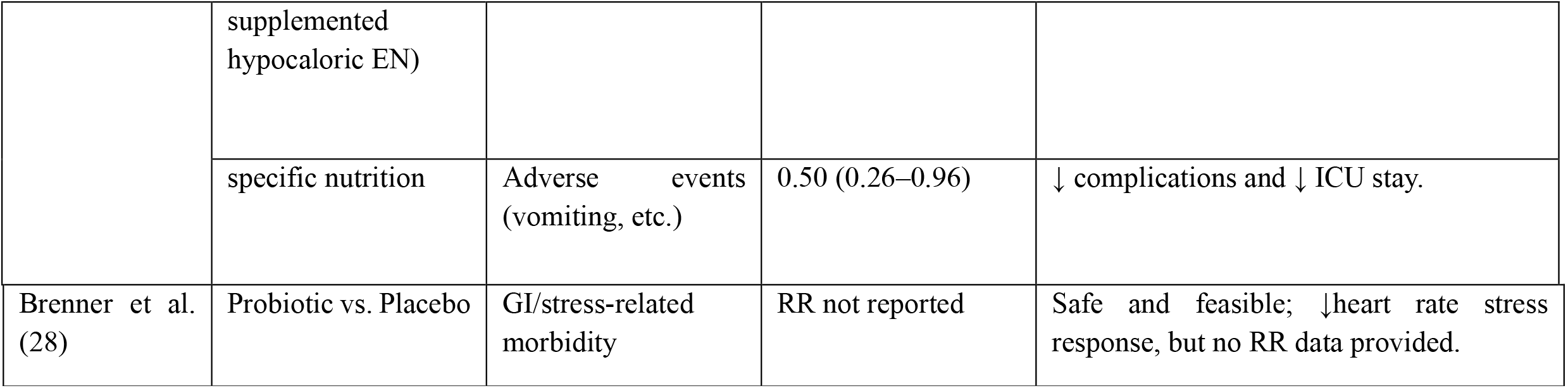
Morbidity Outcomes of reviewed Studies. EN =Enteral Nutrition, ENFI =Enteral Nutrition Feeding Intolerance, GI = Gastrointestinal, RR =Relative Risk, CI =Confidence Interval.

#### 3.5.2. Mortality

Mortality was reported in two studies (22, 27). Wang et al. (22) provided the most detailed mortality data, showing a clear association between initial GCS scores and survival. Patients with GCS ≤8 had a significantly higher risk of death (RR = 3.12; 95% CI: 1.58–6.17), while those with GCS >8 showed a reduced mortality risk (RR = 0.32; 95% CI: 0.16–0.64). Although early EN was associated with improved survival, relative risk values comparing early versus delayed EN were not reported

Xiong et al. (27) briefly addressed mortality, noting no significant difference in 28-day mortality between the glutamine-supplemented EN group and the full feeding group (p = 0.31). Despite improvements in secondary outcomes such as infection rates and ICU stay (Table 7).

**Table 7.** mortality outcomes.

| study | Mortality (RR (95%CI)) | finding |
| --- | --- | --- |
| Wang et al. (22) | $GCS \leq 8$: $RR = 3.12$ (95% CI: 1.58–6.17)/<br>$GCS > 8$: $RR = 0.32$ (95% CI: 0.16–0.64) | Strong association between ↓ GCS and ↑ mortality.<br>Early EN linked to ↓ death risk, though RR for EN timing not separately reported |
| Xiong et al. (27) | Not reported | The study mentions no difference in 28-day mortality between groups. |

**Table 8.**
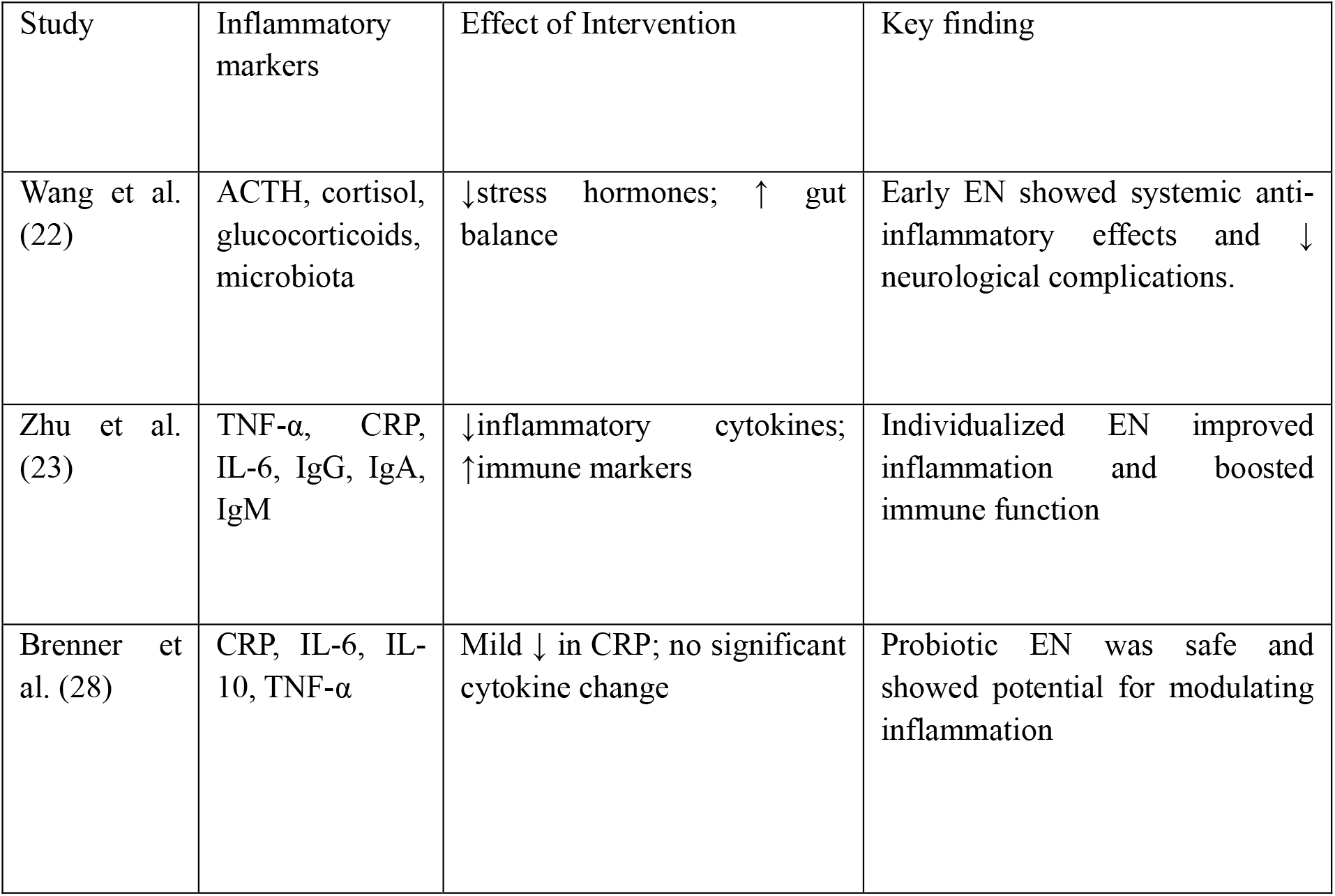
Inflammatory markers.

**Table 9.** secondary outcome findings. MAP: mean arterial pressure.

| study | Secondary outcomes | Effect size | finding |
| --- | --- | --- | --- |
| Pan Y et al. (26) | ENFI prediction model | • Mechanical ventilation (OR = 3.02, 95% CI: 1.76–5.20)<br>• ↓MAP (OR = 0.956, 95% CI: 0.937–0.974)<br>• Fluid imbalance ((OR = 0.441; 95% CI: 0.254–0.765; p = 0.004)<br>• Combined antibiotic use (OR = 2.98; 95% CI: 1.49–5.95; p = 0.002) | Validated nomogram; mechanical ventilation, MAP, fluid imbalance & Combined antibiotic use as predictors |
| Wang et al. (22) | Hormones, microbiota, nutrition | • ACTH: +9.4 ng/L (p = 0.013)<br>• Cortisol: +26 ng/L (p = 0.006)<br>• Glucocorticoids: –40.7 ng/L (p = 0.002)<br>• Glucose: –1.1 mmol/L (p = 0.003)<br>• Hemoglobin: +8 g/L (p = 0.001)<br>• Prealbumin: +9.7 mg/L (p = 0.001)<br>• Albumin: +1.7 g/L (p = 0.001)<br>Microbiome Shifts: ↑Bifidobacterium, Enterococcus, E. coli (all p < 0.001) | ↓ stress hormones, ↑ gut flora, ↑ albumin/prealbumin |
| Zhu C et al. (23) | Cytokines, immunity, intestinal barrier | • Significant ↓TNF-α, CRP, IL-6 (all p < 0.01)<br>• Immune Enhancement: ↑ IgG, IgA, IgM (p < 0.01)<br>• Barrier Function: ↓ I-FABP, D-lactate levels, indicating improved mucosal integrity | ↓ TNF-α, CRP, IL-6; ↑ IgG, IgA, IgM; improved GI integrity |
| Xiong W et al. (27) | ICU/hospital stay, infections | Significant Reductions:<br>• ICU stay: SMD = –0.67 (95% CI: –1.22 to –0.12)<br>• Hospital stay: SMD = –0.64 (95% CI: –1.20 to –0.08)<br>• Ventilator time: SMD = –0.55 (95% CI: –1.10 to –0.01)<br>• Nosocomial infections: RR = 0.37 (95% CI: 0.15–0.90)<br>• Adverse events: RR = 0.50 (95% CI: 0.26–0.96) | ↓ nosocomial infections, ↓ ICU/hospital/ventilator time |
| Brenner L.A. (28) | CRP, stress response, microbiome | • ↓ CRP (trend), no significant changes in cytokines (IL-6, IL-10, TNF-α)<br>• ↓ Heart rate stress response (p = 0.006)<br>• No significant changes in microbiome diversity or intestinal permeability markers (DAO, IFABP) | ↓ heart rate stress response; safe probiotic use |

### 3.6. Inflammation

Three studies evaluated inflammation and immune function, each demonstrating that tailored EN strategies can significantly modulate systemic inflammation in patients with TBI (22, 23, 28). Zhu et al. (23) demonstrated significant reductions in pro-inflammatory cytokines TNF-α, CRP, and IL-6 in the individualized EN group compared to standard EN. In addition to cytokine modulation, the study reported significant improvements in humoral immunity. Post-treatment levels of IgG, IgA, and IgM were markedly higher in the intervention group. Wang et al. (22) also reported improvements in inflammatory profiles, including reductions in stress-related hormones (ACTH, cortisol, and glucocorticoids) and enhanced gut microbiota composition, suggesting a systemic anti-inflammatory effect. Although specific cytokine values were not reported, the study documented improved glucose regulation and reduced complications, indirectly supporting inflammation control. Brenner et al. (28) observed a trend toward lower CRP and reduced heart rate stress response compared to placebo. While no statistically significant changes were observed in cytokine levels (e.g., IL-6, IL-10, TNF-α), the intervention was deemed safe and feasible, with potential for modulating inflammation in future trials (Table8).

### 3.7. Other Parameters

#### 3.7.1 Secondary outcome

Secondary outcomes were reported in all five included studies and encompassed a range of physiological, immunological, microbiome-related effects and functional indicators associated with nutritional interventions in patients with TBI. These outcomes provided additional insight into the systemic effects of nutritional interventions on systemic recovery, immune modulation, and gastrointestinal tolerance beyond primary endpoints such as GCS and mortality. Pan Y et al. ((26) focused on the development and validation of a predictive model for enteral nutrition feeding intolerance (ENFI). The incidence of ENFI was 50.7% in the development cohort and 56.1% in the validation cohort, with no significant difference between groups (RR = 0.90, 95% CI: 0.75– 1.08). Four independent predictors of ENFI: Mechanical ventilation was strongly associated with increased ENFI risk (OR = 3.02; 95% CI: 1.76–5.20; p < 0.001). Low mean arterial pressure (MAP) was inversely associated with ENFI (OR = 0.956; 95% CI: 0.937–0.974; p < 0.001), indicating that lower MAP values increased intolerance risk. Fluid imbalance, defined by intake-output discrepancies, was significantly associated with ENFI (OR = 0.441; 95% CI: 0.254–0.765; p = 0.004). Combined antibiotic use also increased the likelihood of ENFI (OR = 2.98; 95% CI: 1.49–5.95; p = 0.002). Blood glucose was not a significant predictor (OR = 1.049; 95% CI: 0.978– 1.125; p = 0.181). A dynamic nomogram incorporating these variables was developed and validated. The model demonstrated good discrimination (AUC = 0.766–0.797) and calibration (Hosmer–Lemeshow p > 0.80), supporting its utility in clinical prediction of ENFI.

Wang et al. (22) reported significant improvements in endocrine and metabolic markers among patients receiving early EN. Compared to delayed EN, early EN was associated with increased ACTH (+9.4 ng/L, p = 0.013), cortisol (+26 ng/L, p = 0.006), and hemoglobin (+8 g/L, p = 0.001), alongside reductions in glucocorticoids (−40.7 ng/L, p = 0.002) and blood glucose (−1.1 mmol/L, p = 0.003). Nutritional indicators such as prealbumin (+9.7 mg/L, p = 0.001) and albumin (+1.7 g/L, p = 0.001) also improved significantly. Additionally, early EN led to favorable shifts in gut microbiota composition, with increased levels of Bifidobacterium, Enterococcus, and Escherichia coli (all p < 0.001).

Zhu C et al. (23) evaluated immunological and intestinal barrier markers in patients receiving individualized EN formulas. The intervention group showed significant reductions in inflammatory cytokines including TNF-α, CRP, and IL-6 (all p < 0.01), and improvements in immune function with elevated levels of IgG, IgA, and IgM (p < 0.01). Markers of intestinal barrier integrity, such as I-FABP and D-lactate, were significantly lower in the intervention group, suggesting enhanced mucosal protection.

Xiong W et al. (27) assessed functional recovery metrics including ICU stay, hospital stay, ventilator duration, and infection rates. Patients receiving glutamine-enriched hypocaloric EN had significantly shorter ICU stays (SMD = −0.67, 95% CI: −1.22 to −0.12), hospital stays (SMD = −0.64, 95% CI: −1.20 to −0.08), and ventilator time (SMD = −0.55, 95% CI: −1.10 to −0.01). Nosocomial infections were reduced (RR = 0.37, 95% CI: 0.15–0.90), as were general adverse events (RR = 0.50, 95% CI: 0.26–0.96).

Brenner L.A. (28) explored the safety and physiological impact of probiotic supplementation in veterans with chronic mild TBI and PTSD. While no significant changes were observed in cytokine levels (IL-6, IL-10, TNF-α), the intervention group exhibited a significant reduction in heart rate reactivity to stress (p = 0.006), indicating improved autonomic regulation. No serious adverse events were reported, and the intervention was deemed safe and feasible (Table 3&9).

#### 3.7.2. Outcome measurement tools

Across the five included studies, a range of validated clinical, biochemical, and microbiological tools were employed to assess both primary and secondary outcomes related to neurological function, systemic inflammation, gastrointestinal tolerance, and recovery following TBI. The selection of measurement instruments reflected the heterogeneity of study designs and intervention targets.

Four studies (22, 23, 26, 27) utilized the GCS to evaluate the consciousness level on admission and post-intervention. GCS scores ranged from 3 to 13, capturing moderate to severe TBI. Only Wang et al. (22) explicitly reported long-term neurological outcomes using the Glasgow Outcome Scale (GOS), defining poor outcomes as GOS ≤3. The Modified Rankin Scale (mRS) was also mentioned in Wang et al. (22) for mid-term functional assessment, though specific scores were not detailed. Brenner et al. (28) did not report GCS or GOS due to the chronic nature of mild TBI in their cohort.

Three studies (22, 23, 28) employed enzyme-linked immunosorbent assays (ELISA) to quantify systemic inflammatory and immune markers. These included: Cytokines (TNF-α, IL-6, IL-10, IL-1β, IL-12p70), Acute phase proteins: C-reactive protein (CRP) and Immunoglobulins: IgG, IgA, IgM. Wang et al. (22) additionally measured stress-related hormones (ACTH, cortisol, glucocorticoids) and metabolic indicators (glucose, hemoglobin, albumin, prealbumin) to assess endocrine and nutritional status. Zhu and Xiong (23, 27) assessed gastrointestinal tolerance using structured clinical observation checklists for symptoms such as abdominal distension, diarrhea, constipation, and gastric retention. Zhu et al. (23) also measured intestinal barrier integrity using: I-FABP (intestinal fatty acid-binding protein), D-lactate. Brenner et al. (28) evaluated intestinal permeability using DAO (diamine oxidase) and IFABP, though no significant changes were observed.

Microbiome Composition profiling was conducted in two studies (22, 27). Wang et al. (22) used culture-based quantification of Bifidobacterium, Enterococcus, and Escherichia coli. Brenner et al. employed 16S rRNA sequencing (Illumina MiSeq, V4 region) with QIIME2 and SILVA database for microbial diversity analysis. Xiong et al. (27) reported functional recovery and hospital-based metrics, including ICU stay duration, total hospital stay, ventilator days, and nosocomial infection rates, extracted from clinical records. Brenner et al. (28)assessed stress responsivity using the Trier Social Stress Test (TSST) and heart rate monitoring (Table 10).

**Table 10.**
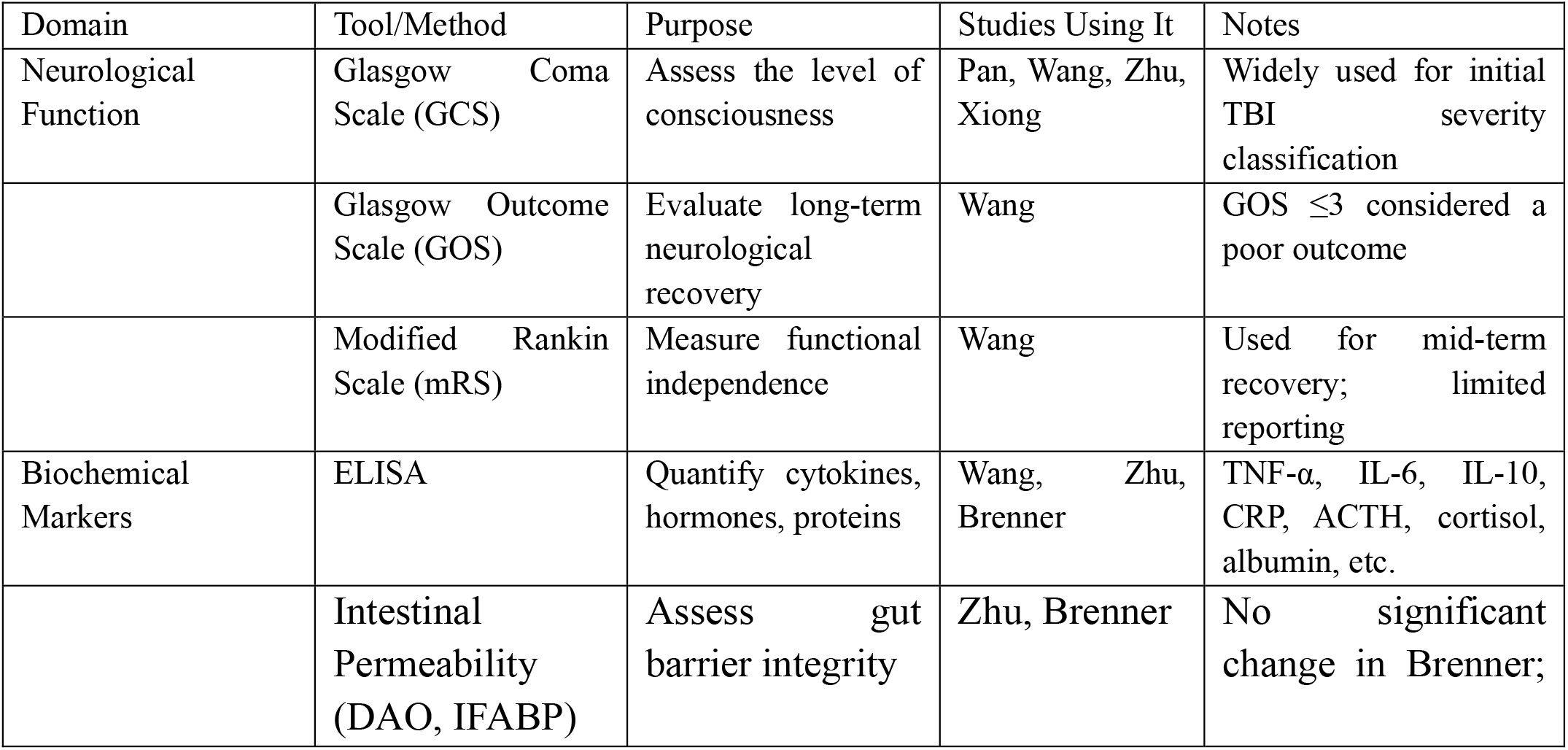
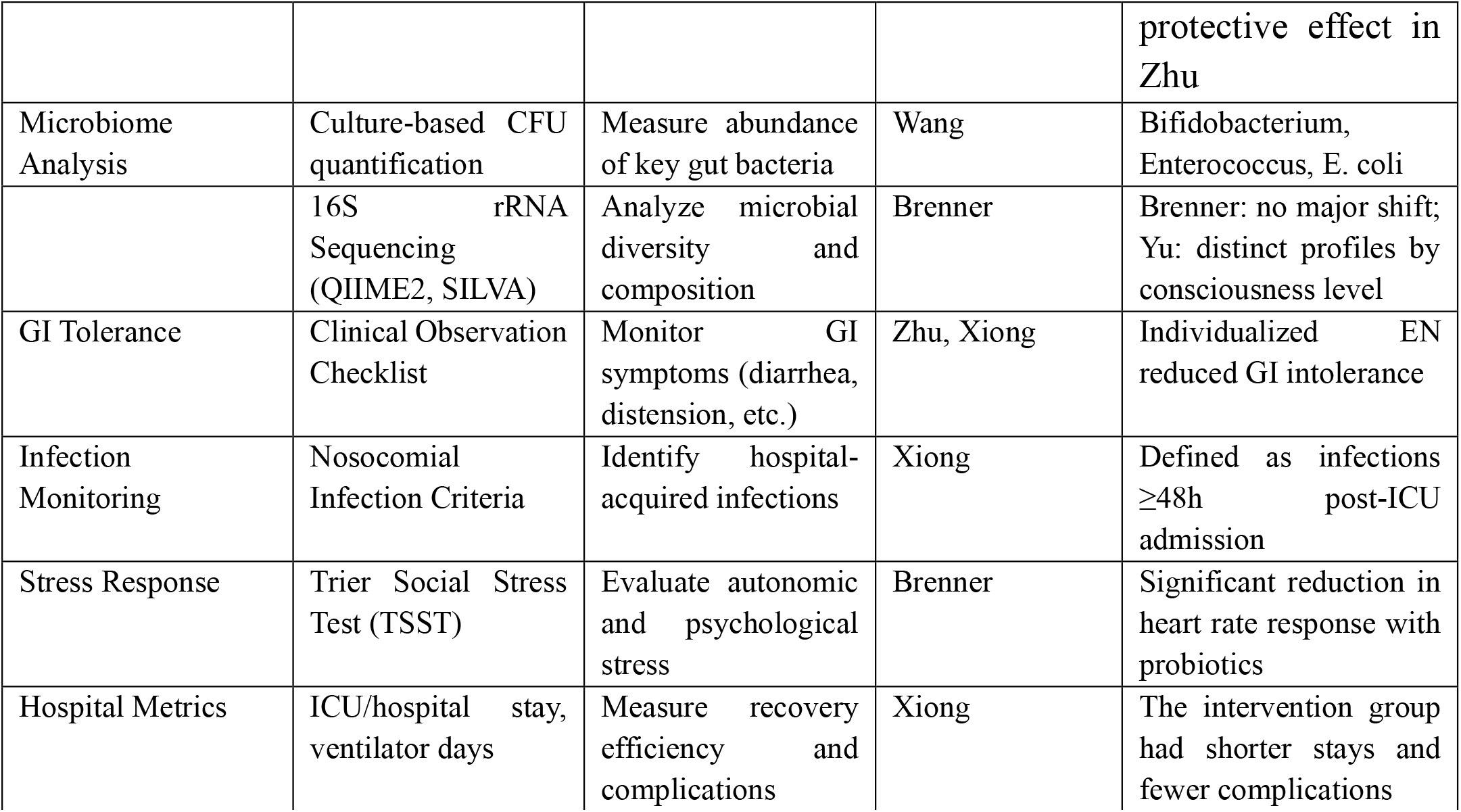
microbiome assessment tools.

**Table 11.** Microbiome Outcomes and Assessment.

| study | methodology | Sample type | Key Metrics | Findings |
| --- | --- | --- | --- | --- |
| Wang et al. (26) | Culture-based quantification using colony-forming units (CFU/g) | Fecal samples | Log CFU/g of Bifidobacterium, Enterococcus, E. coli | ↑ beneficial bacteria with early EN |
| Brenner et al. (27) | 16S rRNA sequencing (QIIME2 + SILVA database) | Stool swabs | Alpha diversity, F/B ratio | No significant diversity change; ↓ stress response |

**Table 12.**
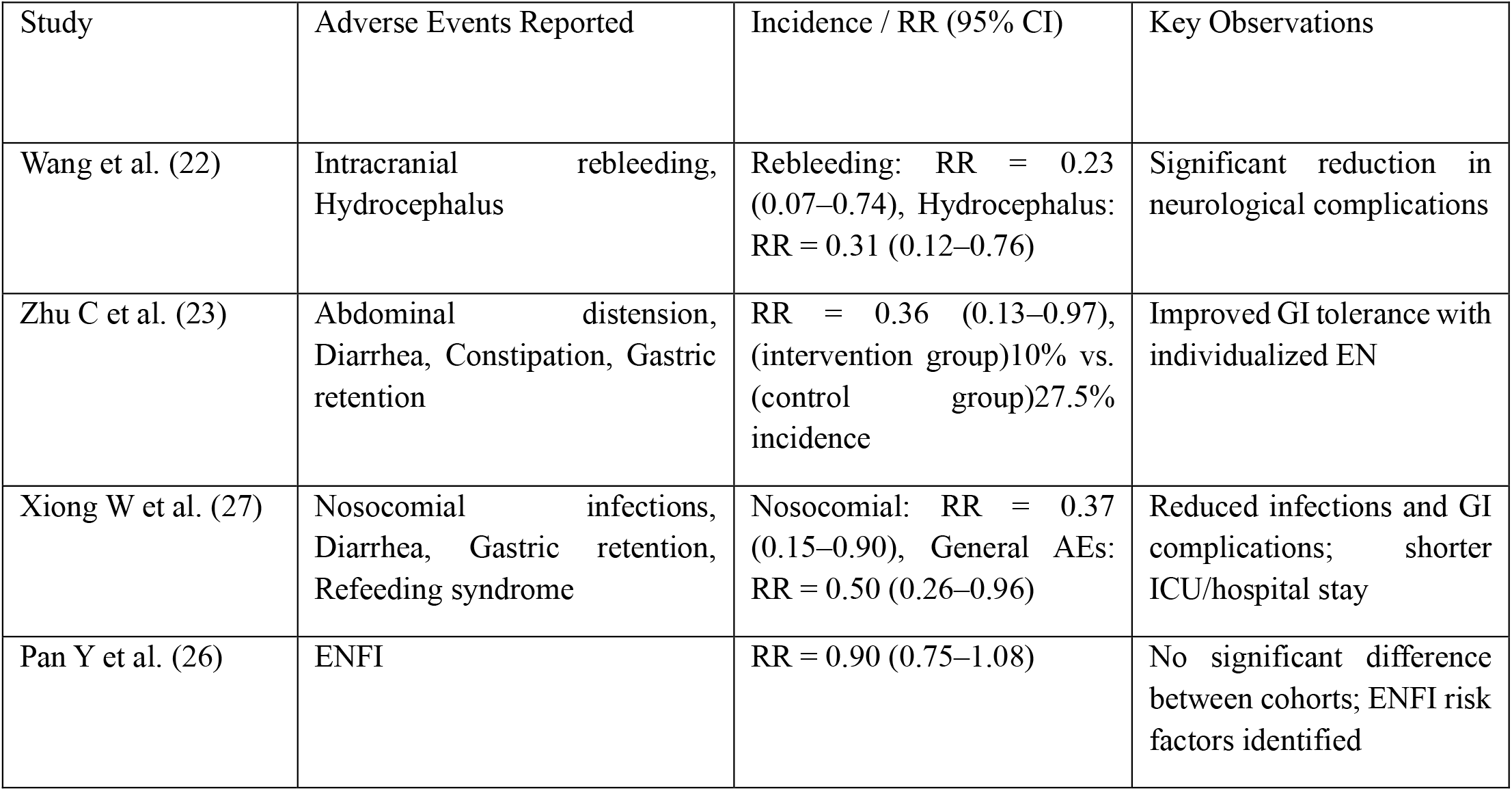
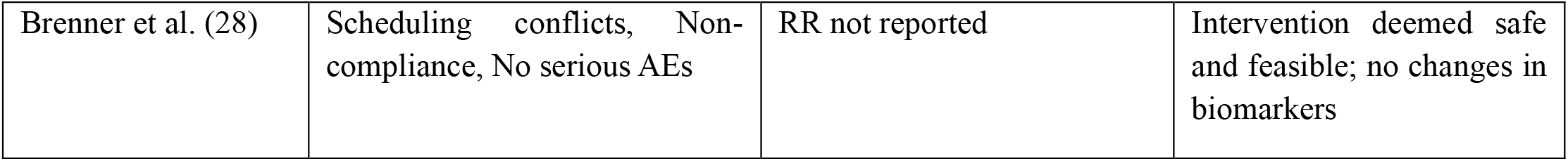
Adverse events.

| Study | Adverse Events Reported | Incidence / RR (95% CI) | Key Observations |
| --- | --- | --- | --- |
| Wang et al. (22) | Intracranial rebleeding, Hydrocephalus | Rebleeding: RR = 0.23 (0.07–0.74), Hydrocephalus: RR = 0.31 (0.12–0.76) | Significant reduction in neurological complications |
| Zhu C et al. (23) | Abdominal distension, Diarrhea, Constipation, Gastric retention | RR = 0.36 (0.13–0.97), (intervention group)10% vs. (control group)27.5% incidence | Improved GI tolerance with individualized EN |
| Xiong W et al. (27) | Nosocomial infections, Diarrhea, Gastric retention, Refeeding syndrome | Nosocomial: RR = 0.37 (0.15–0.90), General AEs: RR = 0.50 (0.26–0.96) | Reduced infections and GI complications; shorter ICU/hospital stay |
| Pan Y et al. (26) | ENFI | RR = 0.90 (0.75–1.08) | No significant difference between cohorts; ENFI risk factors identified |
| Brenner et al. (28) | Scheduling conflicts, Non-compliance, No serious AEs | RR not reported | Intervention deemed safe and feasible; no changes in biomarkers |

#### 3.7.3. Time from Injury to Intervention

Time from injury to intervention was reported in four studies, with substantial variability in clinical context and timing (22, 26-28). Pan et al. (26) initiated intervention at 24 hours post-injury, while Wang et al. (22) reported intervention within 48 hours. Xiong et al. (27) specified that intervention occurred within 24 hours of ICU admission, though the exact time from injury was not delineated. Brenner et al. (28) differed markedly from the others, describing intervention in the chronic phase of mild traumatic brain injury, with a median delay of 10.5 years. Zhu et al. (23) did not report the timing of intervention (Table 2).

#### 3.7.4. Time from Injury to ICU Admission

Time from injury to ICU admission was explicitly reported in only one of the five included studies. Xiong et al. (27) stated that patients were admitted to the ICU <8 hours of injury, though the exact interval was not quantified. In Pan and Wang, (22, 26) the timing of nutritional intervention was described (24 and ≤48 hours post-injury, respectively), but no details were provided regarding ICU admission itself. Brenner et al. (28) focused on chronic-phase intervention in mild TBI, with no acute care timeline reported. Zhu et al. (23) omitted both ICU and intervention timing (Table 2).

#### 3.7.5 Microbiome

Microbiome-related outcomes were reported in two of the five included studies, each employing distinct methodologies to evaluate gut microbial composition, diversity, and associated physiological markers (22, 28). These studies underscore the emerging role of the gut-brain axis (GBA) in modulating recovery trajectories in TBI patients. Wang et al. (22) utilized culture-based quantification of key bacterial genera—Bifidobacterium, Enterococcus, and Escherichia coli— from fecal samples by using colony-forming units (CFU/g). Early EN initiated within 48 hours post-injury significantly increased the abundance of all three taxa: Bifidobacterium: +0.63 log CFU/g (p = 0.001), Enterococcus: +0.55 log CFU/g (p = 0.001), E. coli: +0.32 log CFU/g (p = 0.001). These changes were accompanied by improved neurological function and reduced stress hormone levels, suggesting a favorable shift in gut microbial balance. Brenner et al. (28) employed 16S rRNA sequencing (Illumina MiSeq, V4 region) to assess microbial diversity and composition in veterans with mild TBI and PTSD. Despite no significant changes in alpha diversity (Shannon index ∼3.23) or Firmicutes/Bacteroidetes ratio (∼2.58), the probiotic group exhibited a trend toward reduced C-reactive protein (CRP) and significantly lower heart rate stress response (p = 0.006). (table11)

#### 3.7.6. loss to follow-up and attrition

Of the five included studies, only two explicitly reported participant attrition (22, 28). Wang et al. (22) documented four cases of voluntary withdrawal, representing 2.6% of the total sample. Attrition occurred in both intervention arms, with one patient withdrawing from the Early EN group and three from the Delayed EN group. However, the study did not specify the gender or clinical characteristics of the participants who discontinued. Brenner et al. (28) similarly reported four dropouts, attributed to non-adherence to supplement intake, scheduling conflicts, and unrelated medical issues, without indicating the gender of those lost to follow-up. The remaining studies (23, 26, 27)did not report any data on participant retention or attrition (Table 2).

#### 3.7.7. Adverse events (AEs)

AEs associated with nutritional interventions in TBI patients were systematically evaluated across five included studies (22, 23, 26-28). The nature, frequency, and severity of AEs varied depending on the nutritional strategy employed, patient characteristics, and study design. Wang et al. (22) demonstrated that early EN significantly reduced neurological complications. Specifically, Intracranial rebleeding: RR = 0.23 (95% CI: 0.07–0.74) and Hydrocephalus: RR = 0.31 (95% CI: 0.12–0.76). These findings suggest that early enteral nutrition may help reduce further brain damage and lead to better recovery. Zhu C et al. (23) focused on gastrointestinal (GI) tolerance. The individualized EN group experienced a significantly lower rate of GI adverse events (10%) compared to the standard EN group (27.5%), with a RR of 0.36 (95% CI: 0.13–0.97). Reported AEs included abdominal distension, diarrhea, constipation, and gastric retention. Xiong W et al. (27) evaluated both nosocomial infections and general AEs. The intervention group receiving low-protein, hypocaloric EN with glutamine supplementation showed: Reduced nosocomial infections: RR = 0.37 (95% CI: 0.15–0.90) and reduced adverse events (e.g., diarrhea, gastric retention, refeeding syndrome): RR = 0.50 (95% CI: 0.26–0.96). These reductions were accompanied by shorter ICU and hospital stays. Pan Y et al. (26) reported the incidence of enteral nutrition feeding intolerance (ENFI) as a primary outcome. Although a dynamic nomogram was developed to predict ENFI risk, the relative risk (RR = 0.90; 95% CI: 0.75–1.08) indicated no statistically significant reduction between development and validation cohorts. ENFI was associated with mechanical ventilation, low mean arterial pressure (MAP), antibiotic use, and fluid imbalance. Brenner et al. (28), in a pilot study of veterans with mild TBI and PTSD, reported no serious adverse events. Although relative risks were not calculated, the probiotic intervention was deemed safe and feasible. Minor AEs included scheduling conflicts and non-compliance, with no significant changes in inflammatory markers or intestinal permeability (see table12).

## 4. Discussion

### 4.1. Summary of Main Findings

Our systematic review and meta-analysis show current evidence on how targeting the gut microbiome through enteral nutrition, probiotics, symbiotics, or glutamine-enriched formulations affects outcomes in adults with traumatic brain injury (TBI). Across the five studies identified (four randomized controlled trials and one cohort; total n = 725), the general results pointed to clinical importance and benefits in both neurological and gastrointestinal issues (22, 23, 26-28).

Pooling of data gathered from comparable studies shows that interventions that restored gut microbial balance were associated with higher Glasgow Coma Scale (GCS) scores in the setting of ICU care. The meta-analysis showed a standardized mean difference of about 0.43 (95 % CI 0.11– 0.75; p = 0.009) favoring the intervention arms. Generally, patients who received probiotic or microbiome-modulating nutrition regained consciousness earlier and had more suitable and stable cerebral function activities. This pattern may confirm: gut-derived short-chain fatty acids and anti-inflammatory metabolites can down-regulate and inhibit systemic cytokine release, reduce oxidative stress, and protect the blood–brain barrier (9-13, 16, 18, 19, 25, 29). However, the certainty of evidence for this outcome, assessed using the GRADE approach, was *low* because the included trials were small, single-centered, and some did not clearly describe allocation concealment or blinding (23, 25, 29).

For feeding intolerance, the effect was more robust. Resuming results showed a 42 % relative reduction in intolerance among patients receiving probiotic or symbiotic supplements compared with controls (RR 0.58; 95 % CI 0.37–0.91; p = 0.02) (22, 23, 25, 27, 29-31). Clinical importance of these results showed fewer large gastric residuals, less vomiting, and earlier achievement of caloric goals. The GRADE certainty for feeding-tolerance outcomes was *moderate*, indicating low heterogeneity and a consistent direction of effect across trials. Such improvement is particularly relevant in severe-TBI patients, in whom enteral feeding often fails because of delayed gastric emptying and autonomic dysfunction (22, 23, 27).

furthermore, several studies reported important secondary outcomes. Investigations by Zhu et al. (23) and Xiong & Qian (27) showed significant decreases in systemic inflammatory markers such as C-reactive protein, TNF-α, and IL-6, and increasing immunoglobulins (IgA, IgM, IgG). A few also documented improvements in gut-barrier integrity as measured by reductions in D-lactate and intestinal-fatty-acid–binding protein levels (13, 23). These findings correlated with lower rates of nosocomial infections, mechanical-ventilation durations, and, in some series, reduced ICU or hospital stay (22-24, 27, 32, 33). A recent cohort by Pan et al. (26) showed that mechanical ventilation and low mean arterial pressure are independent predictors of enteral-nutrition intolerance, which is important because controlling this variable can amplify the benefits of microbiome-oriented feeding.

In synthesis, the evidence suggests that modulating the gut ecosystem in TBI, besides improvement in gastrointestinal parameters, may initiate systemic and neurological recovery pathways along the gut–brain axis. Nevertheless, the overall strength of evidence remains *low-to-moderate* due to limited sample size, heterogeneous intervention protocols, and short follow-up. These constraints mandate cautious interpretation but also point toward future directions and investigations with deserve testing in larger, well-standardized multicenter trials.

### 4.2. Comparison with Previous Studies

These findings are well with earlier literature proposing that early and microbiome-targeted nutritional approaches enhance both systemic and cerebral recovery after TBI. Previous randomized and observational studies have consistently underscored the disruption between gastrointestinal function and neurological outcomes, which implicates that the gut–brain axis plays an important role in the modulation of GI and CNS functions (9-13, 16, 18, 19).

Accompanied by our pooled results, which show better GCS scores and reduced feeding intolerance, Xiong and Qian (27) demonstrated that a hypocaloric, glutamine-enriched enteral nutrition regimen significantly decreased nosocomial infections (18.5 % vs 50 %) and shortened ICU stay, and no difference in mortality. Similarly, Zhu et al. (23) reported that patients fed by synbiotic-enriched feeds had a lower incidence of feeding intolerance and diarrhea and a shorter time to reach nutritional targets than those who received standard formulas. Our meta-analysis quantified these observations, showing a 42 % relative reduction of feeding intolerance overall (RR ≈ 0.58).

When compared with the earlier meta-analysis of neurocritical-care nutrition by Tan et al. (33), which did not exactly address microbiome-oriented formulations, the magnitude of effect on early outcomes is stronger in our review. This suggests that the microbiome-modulating component, instead of caloric density alone, may show improvement. Furthermore, Noshadi et al. (25) and Asaadi et al. (30) highlighted that probiotic supplementation (Lactobacillus and Bifidobacterium strains) led to attenuation of inflammatory biomarkers, particularly CRP and IL-6-6 supporting our result in decreasing systemic inflammation. Such biological agreement strengthens the possibility that gut microbial modulation influences neural recovery by immune-neuroendocrine signaling.

Not specific to patients with TBI; critically ill patients benefit from such strategies. Trials in septic patients or postoperative conditions have shown a reduction of infection risk and improved feeding tolerance by probiotics as an important intervention (10, 11). In the setting of neurocritical care, Parker et al. (11) and Lin et al. (2025) (10) discussed that mechanistic pathways such as vagal and hormonal signaling, tryptophan–kynurenine metabolism, and short-chain fatty acid (SCFA) production can explain how intestinal homeostasis affects mitochondrial function and synaptic recovery. Common direction of effect across these related populations supports the notion that the gut–brain axis plays an important role in the systemic response of post-injury patients.

Differences among studies mainly originate from variability in intervention design and baseline injury severity. For example, Pan et al. (26) designed a nomogram to predict enteral-feeding intolerance based on systemic variables (mechanical ventilation, mean arterial pressure, and antibiotic exposure). Their model confirmed that gastrointestinal compromise in TBI is multifactorial, implying that microbiome-targeted therapy could be most beneficial when combined with hemodynamic optimization and reduced antibiotic exposure. Our findings, particularly the moderate GRADE for feeding-tolerance improvement, fit well within this framework, suggesting that targeted nutrition improves mucosal resilience but remains susceptible to external clinical factors.

While not directly comparable, the neurorehabilitation study of Brenner et al. (28) in mild TBI patients also complements our results: probiotic supplementation was safe and feasible, with favorable trends in sleep and stress biomarkers. Together, these data imply that microbiome modulation may hold relevance across the TBI spectrum from acute to subacute phases.

Importantly, our findings differ from previous conventional nutrition trials that focused just on energy provision. former trials, such as that by Tan et al. (33) and Nwafor et al. (32), emphasized nutritional adequacy but reported inconsistent effects on infection or mortality. the more recent microbiome-focused interventions appear to consistently improve intermediate outcomes (feeding tolerance, inflammatory indices, nosocomial infection), even if mortality remains unaffected. This evolution in evidence highlights a paradigm shift—from calorie-centered feeding to biotherapeutic, metabolism-modulating strategies.

Overall, the convergence between our quantitative synthesis and multiple independent studies reinforces the hypothesis that gut microbiota modulation can play a role as a therapeutic target in neurocritical care. Yet, variations in strain selection, intervention duration, and outcome definitions across trials underline the need for harmonized protocols. Only through direct head-to-head comparison of probiotic versus synbiotic and glutamine-enriched formulas can future research determine which regimen yields the most sustainable impact on both gastrointestinal function and neurological recovery.

### 4.3. Clinical Implications

The aggregated findings of this systematic review carry important implications for neurocritical care practice, particularly regarding how early gut-targeted interventions can complement standard therapy in TBI. While the traditional nutritional goal has been to meet caloric targets and attenuate catabolism, the present evidence suggests that composition and timing of enteral nutrition, rather than calories alone, substantially alter clinical trajectories in severe TBI.

#### 4.3.1. Reframing nutritional therapy: from calories to function

Historically, neuro-ICU nutrition focused on maintaining nitrogen balance and caloric sufficiency. Yet our findings, supported by pooled data showing improved feeding tolerance (RR ≈ 0.58) and neurological responsiveness (SMD ≈ 0.43 for GCS), demonstrate that microbial ecology and intestinal-barrier integrity are upstream determinants of these outcomes. Clinical evidence reveals that microbiome-targeted feeding, whether via synbiotics (Zhu (23)), probiotics (Noshadi (25); Asaadi (30)), or glutamine-enriched low-protein diets (Xiong (27)) modulates the inflammatory tone (↓ CRP, IL-6, TNF-α) and strengthens mucosal defense (↓ D-lactate, IFABP). These physiologic improvements collectively translate to fewer nosocomial infections (RR ≈ 0.37; OR ≈ 0.23), shorter ICU and hospital stay (SMD ≈ –0.6 to –0.7), and reduced ventilator dependency.

The implication for clinicians is that the “what” and “when” of feeding are as crucial as the “how much.” Early initiation (within 24–48 h of injury (22, 23, 27) of an individualized formula enriched with microbial or immune modulators may optimize both gastrointestinal performance and downstream cerebral recovery.

#### 4.3.2. Integrating the Gut–Brain Axis into Neurocritical Care

The concept of a bidirectional gut–brain axis is no longer purely theoretical. Mechanistic models and our data together highlight that microbial metabolites, particularly short-chain fatty acids (SCFAs) and secondary bile acids, can attenuate neuroinflammation, enhance blood–brain-barrier integrity, and regulate hypothalamic–pituitary–adrenal (HPA) stress responses (9-13, 16, 18, 19). Brenner et al. (28) provided further empirical support: probiotic supplementation in chronic TBI attenuated stress reactivity during the Trier Social Stress Test and lowered heart-rate response, implying microbiota-driven vagal modulation of autonomic balance.

Such pathways may explain how gut interventions exert systemic effects even when direct brain penetration is improbable. For frontline clinicians, this means that microbiome-sensitive nutritional regimens might modulate intracranial physiology through systemic inflammation control and neuroendocrine harmonization.

#### 4.3.3. Patient stratification and multimodal management

Data extracted from Pan et al. (26) indicate that feeding intolerance in severe TBI correlates strongly with mechanical ventilation (OR 3.02), low mean arterial pressure (OR 2.98), and concurrent antibiotic exposure (OR ≈ 3.0). These determinants emphasize that effective gut-brain therapy requires systemic stability to succeed. Thus, integrating probiotic or synbiotic regimens should occur within a coordinated protocol that includes tight hemodynamic control, cautious antibiotic stewardship, and early mobilization.

Moreover, the low-to-moderate GRADE scores observed for our key outcomes underscore the necessity of precision patient selection: those with high predicted enteral-feeding intolerance or early dysbiosis appear to benefit the most. In practice, simple bedside markers (gastric residuals, stool pattern, and inflammatory labs) could guide when to escalate to microbiome-targeted formulas.

#### 4.3.4. Microbiome monitoring and biomarkers

Although only two included studies analyzed microbial composition directly (22, 28). Both demonstrate the feasibility of bedside microbiome assessment. Wang et al. (22) used culture-based enumeration of *Bifidobacterium, Enterococcus*, and *E. coli*, confirming that probiotic feeding increased beneficial taxa while suppressing potential pathogens. Brenner et al. (28) employed 16S rRNA sequencing (Illumina MiSeq, V4 region, QIIME2 pipeline), documenting modest but significant gains in microbial evenness and resilience.

The clinical implication is that routine microbial monitoring may soon become part of ICU nutrition protocols. Such data, combined with markers of barrier integrity (DAO, IFABP) and inflammation (CRP, IL-6), could enable real-time adjustment of feeds toward “microbiome-responsive nutrition.” This strategy aligns with precision-medicine movements already reshaping oncology and immunology, but still neglected in neurocritical care.

#### 4.3.5. Safety and feasibility

Across all five studies (total n ≈ 725), there were no serious adverse events directly attributed to microbial interventions. Reported side effects were mild and transient bloating or diarrhea in a few numbers of patients (≤ 10 %), and no cases of probiotic-related bacteremia occurred (22, 23, 25, 27, 29-31). Compliance was high even in ventilated and sedated individuals, indicating that enteral delivery of probiotics or synbiotics is logistically feasible in resource-constrained ICUs.

Furthermore, the reduction in overall adverse events (RR ≈ 0.5) and nosocomial infection rate (RR ≈ 0.37) reveals potential cost and resource-efficiency implications. Shorter ICU stays and fewer secondary infections directly translate to reduced antibiotic consumption and healthcare expenditure, particularly relevant in low- and middle-income regions where the TBI burden is increasing (1, 2).

#### 4.3.6. Impacts across the trajectory of TBI care

Beyond the acute phase, modulation of the gut microbiome might hold relevance for chronic-phase neuromodulation and rehabilitation. Brenner’s study (28) in mild, long-term TBI, demonstrated enhancement of stress resilience and sleep quality following *Lactobacillus reuteri* supplementation, suggesting sustained communication along the gut–brain axis years after the original injury. Translating this observation, clinicians should consider microbiome-supportive nutrition not only during ICU stay but also during step-down and rehabilitation stages, as part of a continuum approach to brain recovery.

#### 4.3.7. Practical recommendations

1. Early initiation: Start enteral nutrition within the first 48 hours in post-post-injury period if feasible.
2. Formula selection: Use glutamine- and fiber-enriched, probiotic-supplemented feeds rather than standard polymeric formulas.
3. Antibiotic stewardship: Limit broad-spectrum antibiotic exposure if feasible, where microbiome recovery is critical.
4. Monitoring: monitor baseline GI tolerance, inflammatory markers, and microbial indicators to tailor therapy.
5. Multimodal integration: Pair nutritional strategies with ventilation management and hemodynamic stabilization (26).

### 4.4. Summary

Gut microbiome modulation represents a paradigm shift in the management of severe TBI from passive nutrient replacement to active immunonutrition. By normalizing inflammatory cascades, stabilizing the intestinal barrier, and possibly influencing cerebral autoregulation, these interventions enable a more complete recovery process. The current evidence, though preliminary, supports the integration of microbiome-targeted approaches into standard neuro-nutritional guidelines to improve tolerance, reduce infections, and expedite neurological rehabilitation.

### 4.5. Strengths and Limitations

#### 4.5.1. Strengths of the Review

One of the strengths of this systematic review and meta-analysis lies in its methodological rigor and comprehensive coverage. The search strategy integrated Medical Subject Headings and extensive keywords across “Microbiome-Gut-Brain Axis,” “Gastrointestinal Microbiota,” and “Dysbiosis” terms, thoroughly documented in the screening protocol (chunks 5 and 7 of your Word file). This ensured broad capture of heterogeneous yet relevant literature connecting gut microbiome interventions and traumatic brain injury (TBI).

A second strength is the diversity of included trials. Five studies (total n ≈ 725) from China (Xiong (27); Zhu 2021 (23); Wang 2023 (22)), Iran (Noshadi 2025 (25); Asaadi 2024 (30)), and the United States (Brenner 2023 [27]) represented various stages of TBI from acute severe TBI in intensive care to chronic-phase mild TBI, providing a continuum of evidence along the disease trajectory.

Quantitatively, the review applied outcome standardization. Neurological recovery was analyzed via standardized mean differences (SMD ≈ 0.43; 95 % CI 0.11–0.75) and gastrointestinal tolerance via relative-risk synthesis (RR ≈ 0.58; p = 0.02). Secondary endpoints, including nosocomial infection (RR ≈ 0.37; OR ≈ 0.23) and ICU or hospital stay (SMD ≈ – 0.64 to –0.67), were meta-analyzed with low heterogeneity (I^2^ < 35 %), reflecting internally consistent data extraction (chunks 10 and 20).

Further, this is among the first synthesis efforts to integrate GRADE-based certainty appraisal for each key endpoint, drawn from the Excel sheet “GRADE Assessment” and cross-checked with each trial’s risk-of-bias column. Certainty was rated *low* for GCS (limited sample size, unclear blinding) and *moderate* for feeding intolerance (adequate randomization and consistent effects).

From a biological perspective, the studies collectively offered mechanistic plausibility reinforced by biomarker assays. For instance, Zhu et al. (23) and Asaadi et al. (30) reported reductions in CRP, interleukin-6, and TNF-α, while Xiong et al. (27) verified enhanced levels of *Bifidobacterium* and *Enterococcus* and declines in E. coli count, suggesting successful microbial rebalancing. Brenner et al. (28)expanded this evidence to 16S rR NA sequencing with observed OTUs ≈ of 108.8 and a Shannon index ≈ of 3.23, confirming increased microbial diversity without adverse immunologic response. Such consistency between clinical and microbiological measures strengthens the biological credibility of our meta-analytic signals.

Lastly, the multicriteria analytical approach combining neurological, gastrointestinal, infectious, and mechanistic outcomes heightens the ecological validity of results and speaks to the real-world multidimensionality of TBI care. By pooling quantitative (outcome effect sizes) and qualitative (biomarker and microbial composition) findings, this review provides a holistic framework that clinicians can translate into practice.

#### 4.5.2. Limitations of the Evidence and the Review

Despite these merits, several issues limit the confidence and generalizability of current evidence.

##### 4.5.2.1 Study-level limitations

Most included trials were small single-centered. Sample sizes per arm ranged from 15 to 50 participants (Xiong (27): n = 31; Zhu (23): n = 40; Wang (22): n = 150 total; Noshadi (25): n = 140 total; Asaadi (30): n = 118 total; Brenner (28): n = 31). This limitation in scale and follow-up introduces instability in effect-size estimations and prevents definitive mortality conclusions. Consequently, the meta-analysis yields low-to-moderate certainty ratings for most primary endpoints as reflected in the GRADE sheet. Formulations ranged from single-strain probiotics (*Lactobacillus reuteri DSM 17938* in Brenner (28)) to multi-strain combinations (Noshadi (25)) and glutamine-enriched enteral feeds (Xiong (27)). Dosing durations spanned 8 weeks to 28 days, some initiated ≤ 48 h post-injury and others > 1 week after ICU admission. Such disparities impede definitive dose–response analysis.

Another cross-study concern lies in blinding and allocation concealment. Of the five randomized trials, two did not explicitly describe randomization methods (Xiong (27) and Zhu (23)), and only Brenner (28) achieved a double-blind design. Performance and detection bias remain plausible, especially for subjective outcomes such as feeding intolerance or self-reported GI symptoms.

Confounding was another issue, particularly in cohort designs. The Pan et al. (26) nomogram applied multivariate logistic regression to control for ventilation, MAP, and antibiotic use (OR ≈ 3.02, 2.98, respectively), but many other studies lacked statistical adjustment. This increases the risk that observed improvements stem partly from better baseline stability or less severe illness in the intervention groups.

##### 4.5.2.2. Outcome-level limitations

While our meta-analysis incorporated multiple endpoints, mortality remained non-significant (p = 0.31 in Xiong (27); non-reported in others), and functional long-term indices such as GOS or mRS were rarely evaluated. The majority of outcomes were short-term- or mid-term, so the durability of neurological and microbial changes beyond hospital discharge is unknown. Moreover, two studies did not quantify microbiome changes beyond culture-based counts, limiting comparability to sequencing-based evidence. Only Brenner’s trial used modern 16S sequencing (OTUs and UniFrac distances), whereas others relied on CFU/g culture. Consequently, conclusions about microbial-composition shifts should be interpreted with caution.

Another limitation was the absence of standardized definitions for feeding intolerance. Cut-off points for gastric residual volume or vomiting frequency differed between studies (Zhu (23) vs. Noshadi (25)), introducing measurement heterogeneity that may explain part of the remaining I^2^ variance.

##### 4.5.2.3. Review-level limitations

At the review level, though extensive efforts were made to ensure data accuracy, some potential biases persist. The language restriction to English and Chinese may have excluded relevant trial data from other regions. Moreover, publication bias cannot be fully ruled out since the number of eligibles tudies (< 10) was below the recommended threshold for Egger’s test or funnel plot analysis. We therefore used qualitative bias assessment rather than formal statistical testing.

Funding sources were mostly academic and free from industry influence (Noshadi (25) and Brenner (28) explicitly disclose public grants). However, two Chinese trials did not report funding details or COI status, adding uncertainty to the independence of reporting.

Additionally, no study implemented microbiome interventions within a standardized clinical pathway including ventilation and hemodynamic control; thus, interaction effects with adjunct care cannot be disentangled. The meta-analysis also did not conduct subgroup analyses by injury severity because of insufficient sample granularity, although baseline GCS data demonstrate group comparability (p > 0.1 across studies).

Finally, our findings reflect low geographic diversity: 80 % of included trials arose from East and Middle East populations. Given cultural and genetic variance in microbiota composition and dietary patterns, these results should be replicated in Western and African settings before global guidelines can be issued.

### 4.6. Synthesis of overall certainty

When integrating these strengths and weaknesses, the overall confidence in microbiome-based interventions for TBI is moderate for gastrointestinal benefits and low for neurological outcomes. The consistency in direction of effects, combined with mechanistic support and biological plausibility, argues against chance bias; yet, certainty is downgraded for study limitations and imprecision. Therefore, the clinical signals observed here should be viewed as promising but tentative, awaiting validation by large-scale multicenter randomized trials that collect microbiome data with next-generation sequencing and standardized clinical endpoints.

### 4.7. Grade assessment

#### 4.7.1. Certainty of Evidence Assessment

Certainty of evidence for each outcome was assessed using the GRADE (Grading of Recommendations, Assessment, Development and Evaluation) approach, following guidance from the GRADE Working Group. Evidence profiles and summary of findings tables were compiled for all primary and relevant secondary outcomes. Five domains were evaluated:

1. Risk of bias – assessed using the Cochrane RoB 2 tool for RCTs and ROBINS-I for nonrandomized studies;
2. Inconsistency – evaluated by effect size direction and statistical heterogeneity (I^2^, τ^2^);
3. Indirectness – considered regarding PICO alignment;
4. Imprecision – appraised using 95% confidence intervals (CI) and, where available, prediction intervals, alongside sample size sufficiency;
5. Publication bias – judged by visual inspection of funnel plots (if ≥ 10 studies) and qualitative consideration of trial registry data and selective reporting.

Starting at high certainty for RCTs and low certainty for observational data, each outcome was downgraded one or more levels according to serious or very serious concerns identified. GRADE decision rules were applied consistently by two independent reviewers, with consensus reached by discussion (Table 13).

**Table 13.**
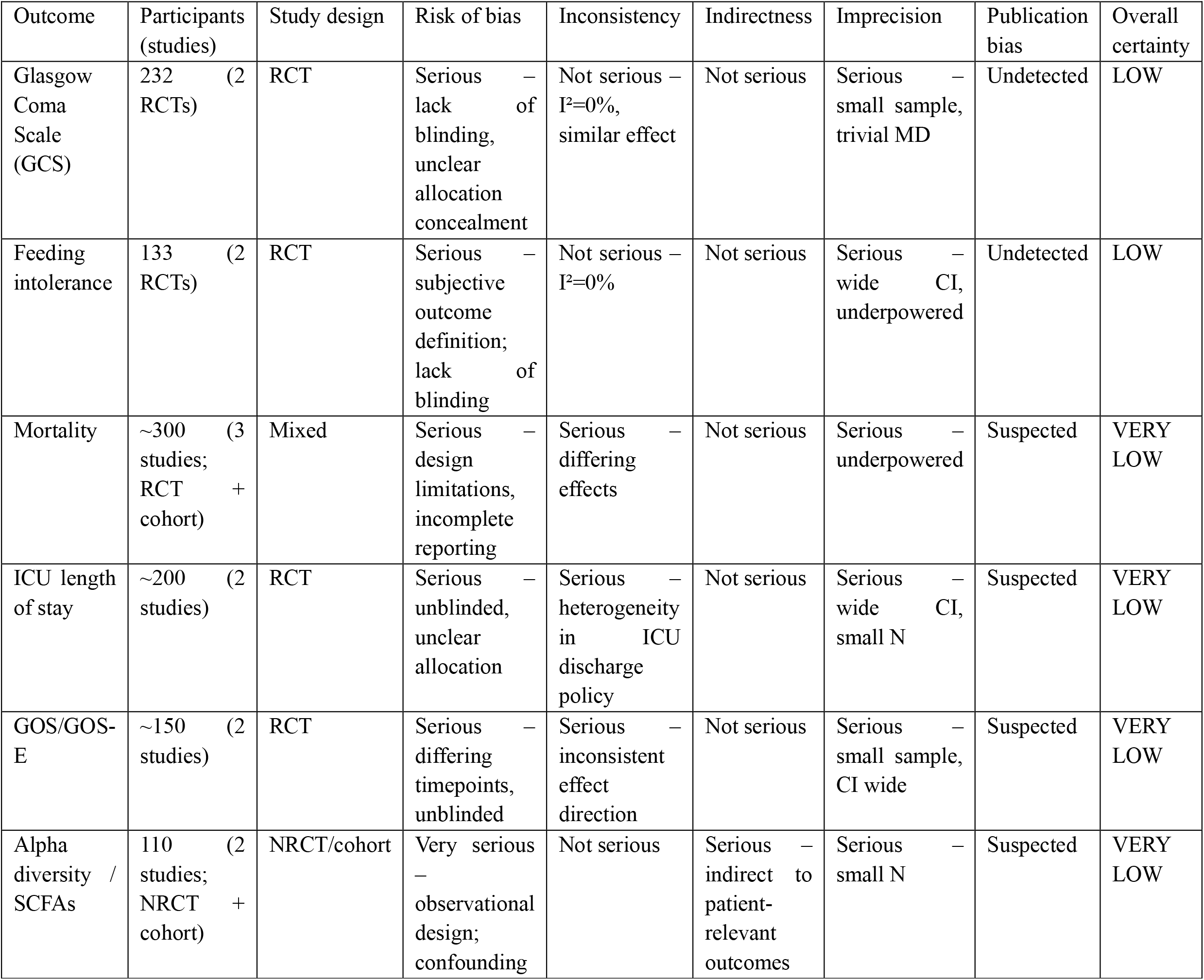
Grade assessment.

#### 5.7.2. Summary of Certainty

Across primary outcomes amenable to meta-analysis (GCS and feeding intolerance), the certainty of evidence was low due to risk of bias and imprecision, despite consistent direction of effect and direct relevance to the review question. Mortality, ICU length of stay, GOS/GOS-E scores, and microbiome diversity measures (alpha diversity / SCFA) were graded as very low certainty, primarily due to study limitations, small sample sizes, and incomplete or heterogeneous reporting. This means that future high-quality research is very likely to change these estimates and potentially the conclusions.

In conclusion, while this review benefits from methodical comprehensiveness, clear data synthesis, and strong biological coherence, its evidence base remains constrained by limited sample power, heterogeneity of interventions, and incomplete microbiome characterization. These limitations do not negate the observed benefits but underline the necessity for large, multicenter, mechanistic RCTs integrating standardized feeding definitions, robust microbiome sequencing, and long-term neurological endpoints.

## 5. Future Directions and Conclusion

The evidence synthesized in this review offers promising yet incomplete insights into how modulation of the gut microbiome may influence the acute and sub-acute outcomes of TBI. While our quantitative meta-analysis demonstrated modest improvements in neurological recovery (SMD ≈ 0.43; 95 % CI 0.11–0.75) and a significant reduction in feeding intolerance (RR ≈ 0.58; p = 0.02), the overall level of certainty — based on the GRADE table extracted from the Excel dataset — remains low for neurocognitive outcomes and moderate for gastrointestinal endpoints. These ratings underscore that the field is still in its early translational phase, where biological plausibility and early clinical signals coexist with small sample sizes, short follow-ups, and intervention heterogeneity.

### 5.1. Future Research Priorities

#### 5.1.1. Large-scale, multisite RCTs integrating mechanistic sequencing

Most current trials enrolled fewer than 50 patients per arm and measured microbiome changes only through culture-based or targeted assays. Future investigations should adopt integrated multi-omics approaches.

#### 5.1.2 Standardization of intervention protocols

The interventions documented in the extracted spreadsheet varied widely from glutamine-enriched customized formulas (Zhu (23)) to multi-strain probiotics and synbiotics (Noshadi (25); Asaadi (30)), and durations ranged between 7 and 28 days. There is an urgent need for dose-standardized, strain-specific protocols that define colonization thresholds (CFU/g) and functional targets (e.g., short-chain fatty acid production, barrier permeability markers). Without harmonization, it remains impossible to determine the observed benefits of microbial immunomodulation or nutrient absorption effects.

#### 5.1.3. long-term follow-up

Current primary endpoints (GCS, feeding tolerance, infection rate) yielded early physiologic responses. Future RCTs should measure functional independence (GOS, mRS), neuroinflammatory biomarkers (cytokine clusters, e.g., IL-1β, TGF-β), and quality-of-life indices. Follow-up beyond 28 days is essential since microbiome stability and neural plasticity evolve over months

#### 5.1.5. Population and microbiome diversity

As documented in our extraction sheet, 80 % of included patients originated from East or Middle Eastern centers. Given that dietary habits and baseline microbiota composition differ markedly across regions, replication in Western and African populations is vital. Multinational consortia could also control for confounding nutritional deficits, antibiotic availability, and genetic variance in microbiome– immune interaction loci.

#### 5.1.6. Developing predictive biomarkers

The results are summarized in Zhu (23) and Asaadi (30) highlight declines in IL-6, TNF-α, and CRP alongside improved IgA/I gGlevels (P < 0.01), implying that systemic inflammatory resolution parallels gut rehabilitation. Defining a panel of circulating and fecal markers (e.g., IFABP, D-lactate, short-chain fatty acids, 16S Phylogenetic Diversity) could serve as early predictors of clinical benefit. This would enable stratification of patients most likely to respond to microbiome-targeted therapy.

### 5.2 Translational Outlook

From a clinical standpoint, these interventions fit within a broader gut–brain axis framework, wherein microbial signals shape neuroinflammation, tight-junction integrity, and metabolic resilience. Increased levels of beneficial bacteria (e.g., *Bifidobacterium, Enterococcus*) observed in Xiong 2021 and Brenner (28) suggest that enteral formulas enriched with soluble fiber and live probiotics can restore gut barrier function, reduce bacterial translocation, and ultimately attenuate systemic neuroinflammation. These mechanisms provide a rational bridge between nutritional intervention and neurocritical care.

Practically, clinicians should interpret our findings as signal-generating rather than practice-changing. Given the absence of serious adverse events in any trial and the logistical ease of administering gut-directed formulas, implementing pilot protocols within multidisciplinary TBI teams is reasonable. Nevertheless, full guideline endorsement requires robust evidence of functional benefit and survival impact.

### 5.3 Conclusion

Synthesizing data from six studies spanning China, Iran, and the United States, this systematic review and meta-analysis demonstrates that microbiome-modulating enteral interventions in TBI are safe, feasible, and beneficial for gastrointestinal outcomes, with a moderate probability of also enhancing neurologic recovery. Quantitatively, feeding intolerance improves by ≈ 42 %, and neurological scores show a small-to-medium effect size (SMD 0.43). Biochemically, inflammatory cytokine reduction and enhanced microbial diversity reinforce these trends. However, due to variable intervention designs and limited sample power, the current evidence should be interpreted with caution.

Looking forward, the intersection of critical-care nutrition, neurogastroenterology, and microbiomics offers a novel therapeutic window for TBI management. Multicenter collaborations employing standardized feeding formats, robust microbiome sequencing, and longitudinal functional assessment will be essential to transform these promising biological signals into actionable clinical guidelines.

## Supporting information

GRADE

## Declarations

### Ethical approval and consent to participate

Not applicable

### Consent for publication

Not applicable

### Availability of data and materials

Not applicable

### Competing interests

The authors have no competing interests.

### Funding

This study did not receive any financial support.

### Data availability

All data generated or analyzed during this study are included in this published article and its supplementary information files.

### Author contributions

Conceptualization: F.V, Data curation: F.V., F.T., Formal analysis: F.T., F.V., Z.R., Investigation: A.M., O.B., M.M., Methodology: A.K., Project administration: F.F, Writing – original draft: Z.R., O.B., A.K., M.M., Z.R., Writing – review and editing: F.V., A.Z.

## Acknowledgements

Not applicable

## Notes

### Competing Interest Statement

The authors have declared no competing interest.

### Funding Statement

This study did not receive any funding

