## Supplementary material for "Modulation of Gut Microbiota and Gut-Brain Axis as a Therapeutic approach in Traumatic brain injury: Implications for Neurological Outcomes": GRADE

SUPPLEMENTORY - Methods for Certainty of Evidence Assessment

Certainty of evidence was assessed for all primary and secondary outcomes according to the GRADE Working Group methodology. All randomized controlled trials (RCTs) began at ‘high certainty’, and observational studies at ‘low certainty’, with downgrading based on five domains:

1. Risk of bias – evaluated using the Cochrane RoB 2 tool for RCTs and ROBINS-I for non-randomized or cohort data;
2. Inconsistency – assessed through effect size direction and statistical heterogeneity (I², τ², χ²);
3. Indirectness – judged against the PICO framework for alignment between population, intervention, comparator, and outcome;
4. Imprecision – based on width of 95% confidence and prediction intervals, and optimal information size;
5. Publication bias – appraised using funnel plot inspection (where ≥10 studies were available) and registry searches.

GRADE Evidence Profile Table

| Outcome | Participants (studies) | Study design | Risk of bias | Inconsistency | Indirectness | Imprecision | Publication bias | Overall certainty |
| --- | --- | --- | --- | --- | --- | --- | --- | --- |
| Glasgow Coma Scale (GCS) | 232 (2 RCTs) | RCT | Serious – lack of blinding, unclear allocation concealment | Not serious – I²=0%, similar effect | Not serious | Serious – small sample, trivial MD | Undetected | LOW |
| Feeding intolerance | 133 (2 RCTs) | RCT | Serious – subjective outcome definition; lack of blinding | Not serious – I²=0% | Not serious | Serious – wide CI, underpowered | Undetected | LOW |
| Mortality | ~300 (3 studies; RCT + cohort) | Mixed | Serious – design limitations, incomplete reporting | Serious – differing effects | Not serious | Serious – underpowered | Suspected | VERY LOW |
| ICU length of stay | ~200 (2 studies) | RCT | Serious – unblinded, unclear allocation | Serious – heterogeneity in ICU discharge policy | Not serious | Serious – wide CI, small N | Suspected | VERY LOW |
| GOS/GOS-E | ~150 (2 studies) | RCT | Serious – differing timepoints, unblinded | Serious – inconsistent effect direction | Not serious | Serious – small sample, CI wide | Suspected | VERY LOW |
| Alpha diversity / SCFAs | 110 (2 studies; NRCT + cohort) | NRCT/cohort | Very serious – observational design; confounding | Not serious | Serious – indirect to patient-relevant outcomes | Serious – small N | Suspected | VERY LOW |

**Detailed Justifications per Domain**

**Risk of bias:**

- GCS: Both RCTs lacked blinding; allocation concealment unclear in Zhu 2021. Single-center designs.
- Feeding intolerance: Similar limitations plus non-standardized, subjective intolerance measures in Zhu 2021 and Xiong 2021.
- Mortality: mix of study designs; unclear reporting of attrition.
- ICU LOS and GOS/GOS-E: absence of blinding; discharge policies varying.
- Microbiome: uncontrolled observational designs; strong risk of confounding.

**Inconsistency:**

- No downgrade for GCS & feeding intolerance: consistent direction, I²=0%.
- Downgrade mortality, ICU LOS, GOS due to divergent effect size directions.

**Indirectness:**

- Most clinical endpoints align with the PICO; microbiome endpoints indirect to patient-relevant clinical recovery.

**Imprecision:**

- Downgrade for all due to small number of studies and wide confidence intervals; both GCS and feeding intolerance borderline underpowered for clinical decision-making.

**Publication bias:**

- Cannot be excluded; only 2–3 small trials per outcome, many from single-country contexts.
- Across outcomes eligible for synthesis:
- **GCS:** Pooled SMD = –0.15 (95% CI –0.29 to –0.02; p≈0.025; I²=0%, τ²=0.00) from Wang 2023 and Zhu 2021 shows trivial, statistically detectable but clinically unimportant reduction in GCS in intervention arms over 14 days. Both trials high risk for bias due to absence of blinding.
- **Feeding intolerance:** RR = 0.45 (95% CI 0.10 to 2.06; p≈0.095; I²=0%, PI=0.02–12.80); consistent reduction in intolerance in Zhu 2021 and Xiong 2021 but nonsignificant pooled effect due to imprecision.
- **Mortality:** Heterogeneous both in direction and timing of assessment; incomplete event data from some studies; very low certainty.
- **ICU LOS:** High variability in discharge criteria; insufficient harmonisation of outcome measurement; very low certainty.
- **GOS/GOS-E:** Timepoint mismatch prevents confident pooling; inconsistent effect direction; very low certainty.
- **Alpha diversity / SCFAs:** Microbiome alterations significant within studies (Yu 2022) but indirect to neurological function; observational design limits certainty.

Given downgrades primarily for risk of bias and imprecision and, for several outcomes, inconsistency, the certainty of evidence is **low** for GCS and feeding intolerance and **very low** for all other endpoints. Interpretation in clinical practice must be cautious, and definitive guidance awaits larger, multicenter, methodologically rigorous trials.
